# Which assessments are used to analyze neuromuscular control by electromyography after an anterior cruciate ligament injury to determine readiness to return to sports? A systematic review

**DOI:** 10.1101/2020.03.05.20031617

**Authors:** Angela Blasimann, Irene Koenig, Isabel Baert, Heiner Baur, Dirk Vissers

## Abstract

**Background:** Adequate neuromuscular control of the knee could be one element to prevent secondary injuries after an anterior cruciate ligament (ACL) injury. To assess neuromuscular control in terms of time, amplitude and activity, electromyography (EMG) is used. However, it is unclear which assessments using EMG could be used for a safe return to sports (RTS). Therefore, we aimed to summarize EMG-related assessments for neuromuscular control of the knee in adult patients after an ACL injury to decide upon readiness for RTS.

**Methods:** This systematic review followed guidelines of Preferred Reporting of Items for Systematic Reviews and Meta-Analyses (PRISMA) and Cochrane recommendations. MEDLINE/PubMed, EMBASE, CINAHL, Cochrane Library, Physiotherapy Evidence Database (PEDro), SPORTDiscus and the Web of Science were searched from inception to March 2019 and updated in November 2020. Studies identifying electromyographic assessments for neuromuscular control during dynamic tasks in adult, physically active patients with an anterior cruciate ligament injury were eligible and qualitatively synthesized. Two independent reviewers used a modified Downs and Black checklist to assess risk of bias of included studies.

**Results:** From initially 1388 hits, 38 mainly cross-sectional, case-controlled studies were included for qualitative analysis. Most studies provided EMG outcomes of thigh muscles during jumping, running or squatting. Outcomes measures described neuromuscular control of the knee in domains of time, amplitude or activity. Risk of bias was medium to high due to an unclear description of participants and prior interventions, confounding factors and incompletely reported results.

**Conclusions:** Despite a wide range of EMG outcome measures for neuromuscular control, none was used to decide upon return to sports in these patients. Additional studies are needed to define readiness towards RTS by assessing neuromuscular control in adult ACL patients with EMG. Further research should aim at finding reliable and valid, EMG-related variables to be used as diagnostic tool for neuromuscular control. Moreover, future studies should aim at more homogenous groups including adequately matched healthy subjects, evaluate gender separately and use sport-specific tasks.

**Registration:** The protocol for this systematic review was indexed beforehand in the International Prospective Register of Systematic Reviews (PROSPERO) and registered as CRD42019122188.

## Background

Anterior cruciate ligament (ACL) injuries happen quite frequently and concern athletes (0.15 injuries per 1000 athletic exposures (AEs)) but also the active part of the general population (1, 2). Most ACL injuries are due to a non-contact, multiplane mechanism (3) and may lead to instability, secondary meniscal injury or even knee osteoarthritis in the long run (4). Consequently, this injury means several months or even years of physical impairment with wide consequences for the patients concerning return to work, return to activity or return to sport (RTS). RTS rates between 63 and 97% are reported for patients after ACLR (5, 6). Most elite athletes return to sports return earlier than non-elite athletes (5), on average within 12 months (6). However, it remains unclear whether this approach is safe (6), omitting further injury, respectively. Athletes after ACLR returning to high-demanding sports (including jumping, pivoting and hard cutting) show a more than fourfold increase in reinjury rates over two years (7). More than 5% of athletes with an ACLR sustain a re-rupture of the graft (6, 8) in the ipsilateral knee after RTS. The risk for an ACL tear in the contralateral knee is as double as high (11.8%) even five years or longer after an ACLR (8). Overall, the recurrence rates even after successful ACLR and subsequent rehabilitation are high (29.5% or 1.82/1000 AEs), with a tear of the ACL graft (9.0%), an ACL injury of the opposite leg (20.5%), muscle injuries on the ipsilateral side or even bilateral consequences (9, 10).

It is known that ipsilateral deficits in clinical knee function and knee laxity persist even years after ACLR (11, 12). ACL patients show altered kinematics and kinetics (13) and different neuromuscular strategies during walking (14), not only in the injured limb but also in the non-affected side (13, 15). These changes are referred to neuromuscular adaptations due to altered sensorimotor control (16) and are caused by altered afferent inputs to the central nervous system due to the loss of the mechanoreceptors of the native (original) ACL (17). Current literature regarding in ACL patients emphasises the importance of understanding consequences of ACL injury regarding neuromuscular control and kinematics (18–20). To describe neuromuscular control in terms of simultaneously activated agonist/antagonist muscle pairs, generalized knee muscle co-contraction parameters are used (21, 22).

In daily clinical practice, physical performance tests batteries including jumps and tests of muscle function (23) are often used to assess neuromuscular control for RTS. However, there is only limited evidence that passing RTS test batteries – interpreted as having achieved adequate levels of mobility, stability, strength, balance, and neuromuscular control for RTS - reduces the risk for a second ACL injury (24). Moreover, it remains unclear which measures should be used to bring athletes safely back to RTS with a low risk of re-injury (25). In conclusion, the currently suggested RTS criteria do not seem to be adequate to assess neuromuscular control of the knee joint to judge upon a safe RTS or even competition. Therefore, meaningful, reliable, valid and accurate diagnostic tools for patients with an ACL injury (either treated surgically or conservatively) are needed and may aid clinical decision-making towards a safe RTS following ACLR. Objective measurements of neuromuscular control should include electromyography (EMG) of involved muscles to judge upon quantity, quality and timing of voluntary activation and reflex activity (13, 20, 26). However, up to date it is unclear which EMG-related measurements for neuromuscular control are used in patients with an ACL injury to decide upon a safe RTS.

### Objectives

The first objective of this systematic review was therefore to summarize the scientific literature regarding EMG-related assessments for neuromuscular control in adult, physically active patients with an ACL injury (either treated surgically or conservatively) during functional tasks. The second aim was to analyze whether these assessments for neuromuscular control were used to decide upon readiness for RTS in these patients.

## Methods

This systematic review was planned, conducted and analyzed according to the guidelines of Preferred Reporting of Items for Systematic Reviews and Meta-Analyses (PRISMA) (27) and followed the recommendations of Cochrane group (28).

The protocol for this systematic review was indexed beforehand in the International Prospective Register of Systematic Reviews (PROSPERO) and got the registration number CRD42019122188.

### Eligibility criteria

To define the relevant key words for the literature search, the Participants-Intervention-Control-Outcome-Study design (PICOS) scheme was used as follows (Table 1):

**Table 1:**
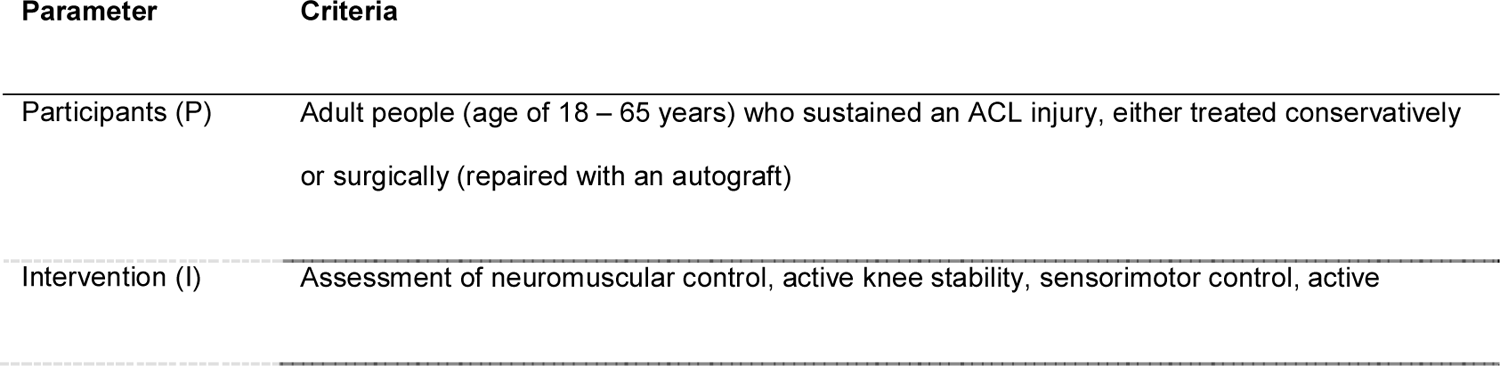
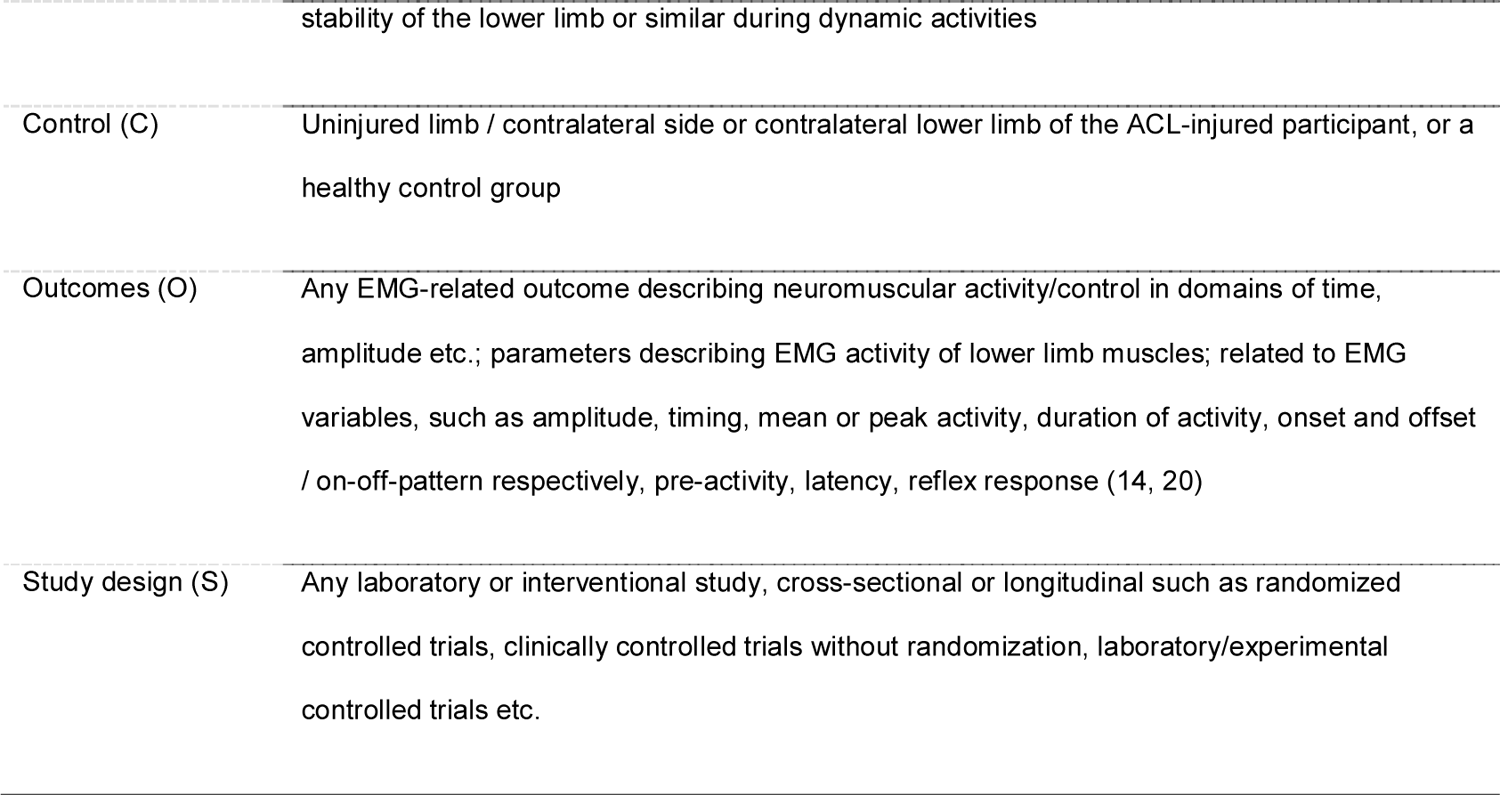
Overview of PICOS criteria for key word definitions

Studies were considered eligible for this systematic review if they met the following inclusion criteria: Study participants – either females, males or both - had to be 18 years or older, suffer from an ACL injury - either treated conservatively or surgically - with a time since injury/surgery of six months at least, be athletes or physically active people who participate in sports activities on a regular basis (as defined by each study, e.g. Tegner Activity Score (TAS) ≥ 3) to get data to decide upon RTS. Moreover, included studies had to have used active or functional tasks such as walking, stair climbing or jumps, applied assessments for neuromuscular control of lower limb muscles using EMG, be original articles published in peer-reviewed, scientific journals in English, German, French, Italian or Dutch, and available as full texts. Exclusion criteria were model-driven approaches, animals or cadavers, comparisons of surgical techniques, passive or non-functional tasks (such as isokinetic measurements for strength and isometric muscle activity), editorials, conference abstracts, book chapters, theses, systematic reviews and meta-analyses.

### Data sources

The search was effectuated from inception until March 2019 and updated in November 27^th^, 2020 in the electronic databases MEDLINE/PubMed, EMBASE, CINAHL, Cochrane Library, Physiotherapy Evidence Database (PEDro), SPORTDiscus and in the Web of Science. To ensure new articles matching the search terms, e-mail alerts were established from each of the databases if possible (29). Furthermore, a hand search was done using the reference lists of included articles to identify additional and potentially eligible articles that had been missed in the electronic database searches. The hits from these two additional sources were also screened for eligibility applying the same criteria as for the articles from the database search.

### Search strategy

In all sources, the advanced search mode was used if available. A search matrix combining relevant keywords (if possible MeSH-terms) with the Boolean operators AND and OR was used and customized for searches in all databases if necessary (see Appendix A): “anterior cruciate ligament/anterior cranial cruciate ligament/ACL”; “anterior cruciate ligament injuries/strains and sprains/rupture/tear/injury/deficiency”; “anterior ligament reconstruction/anterior cruciate ligament/surgery/reconstructive surgical procedures/orthopedic procedure/orthopedic procedure/tendon graft/tendon transfer/conservative treatment/non-surgical/rehabilitation/physical therapy modalities/physiotherapy/kinesiotherapy/ exercise/ instruction/resistance training/neuromuscular training/postoperative care”; “neuromuscular control/neuromuscular activity/sensorimotor control/muscle activity/active stability”; “electromyography/EMG/electromyogram/amplitude/timing/mean activity/peak activity/duration of activity/onset/offset/on-off-pattern/pre-activity/latency/reflex response”. In the updated search, articles were filtered by date of publication, with the aim of including only those published between March 2019 and November 2020.

### Study selection

All hits obtained by the database searches were downloaded to the Rayyan reference management platform (30) and inserted into EndNote (Clarivate Analytics, Philadelphia, USA). Prior to screening, duplicates were removed. Two authors (AB and IK) independently screened title and abstract of the records, one by using the software EndNote (Clarivate Analytics, Philadelphia, USA) and the other with the help of the free software “rayyan” (30). After screening, full texts of relevant hits were read by the two authors (IK, AB) to decide upon in- or exclusion. If their decisions did not match, discussion took place until consensus was achieved. If consensus would not have been reached, a third author (IB or HB) would have finally decided upon in- or exclusion of the record in question; however, this was not necessary.

### Data collection process and data extraction

After final decision of all studies, data extraction for each eligible study was performed by the first author (AB) with a predefined Microsoft® Excel (Microsoft Corporation, Redmond WA, USA) spreadsheet as piloted form. The first author (AB) extracted necessary information from each article describing the study design, groups measured and their characteristics, the tasks to be fulfilled by all participants, and all EMG-related assessments or methods used to evaluate neuromuscular control. Furthermore, the chosen assessment for neuromuscular control were judged whether they were used to clear the participants for RTS. The second author (IK) checked the extracted data at random. As all included studies provided enough information to be qualitatively analyzed, it was not necessary to contact corresponding authors for obtaining or confirming data.

### Assessment of risk of bias in included studies

The risk of bias of all the included articles was independently assessed by two raters (AB, IK) by using the Downs and Black checklist (31) in a modified form (29, 32). The following categories were evaluated: (i) reporting bias: objectives/hypothesis, main outcomes, patients’ characteristics, interventions, principal confounders, main findings, estimates of random variability, actual probability values; (ii) external validity bias: study subjects/staff/places/facilities representative; (iii) internal validity bias: blinding subjects/assessors, data dredging present, different lengths of follow-up/same time period between intervention and outcome for cases and controls, statistical tests/main outcome measures appropriate; (iv) selection bias: patients and controls from same population and over same period of time, randomization, allocation concealed, adjustments for confounding, loss to follow-up; and (v) power analysis (see Appendix B). Each question of the categories was scored with 1 or 2 points if the criterium was fulfilled (answer “yes”), zero points if the answer was “no”, “not fulfilled” and an “X” if the criterium was not applicable, e.g. randomization for a case-control or cross-sectional study, “IC” for intrasubject comparison, respectively.

For this systematic review, studies with a total score of 17 or above out of 25 (more than 2/3 of the maximum total score) were considered as being of high methodological quality, showing a “low” risk of bias respectively (29). Studies which reached 13 to 16 points (more than 50% of the maximum total score) were rated as being of “medium” quality, and total scores below 13 were rated as being of low methodological quality, “high” risk of bias respectively. As the aim of this systematic review was to summarize the applied measures for neuromuscular control, the methodological quality of the included studies was of secondary interest. Therefore, no study was excluded due to a low total score in the risk of bias assessment.

## Results

### Study selection

Hits from the first and the updated database search including e-mail alerts and hand search were screened for duplicates. After applying in- and exclusion criteria according to PRISMA flowchart (27), a total of 38 articles involving 1236 subjects – 809 participants with ACLR or ACL deficiency and 427 healthy controls - could be used for qualitative analysis. Reasons for exclusion were participants younger than 18 years, not able to achieve RTS, time since injury or surgery less than six months, static or non-functional task, study design (e.g. systematic review, study protocol), unclear or inadequate outcome, healthy participants or without ACL injury. Included studies had mainly a cross-sectional, case-controlled study design. Details about every step of the search are illustrated in the following flowchart (Figure 1).

**Figure 1:**
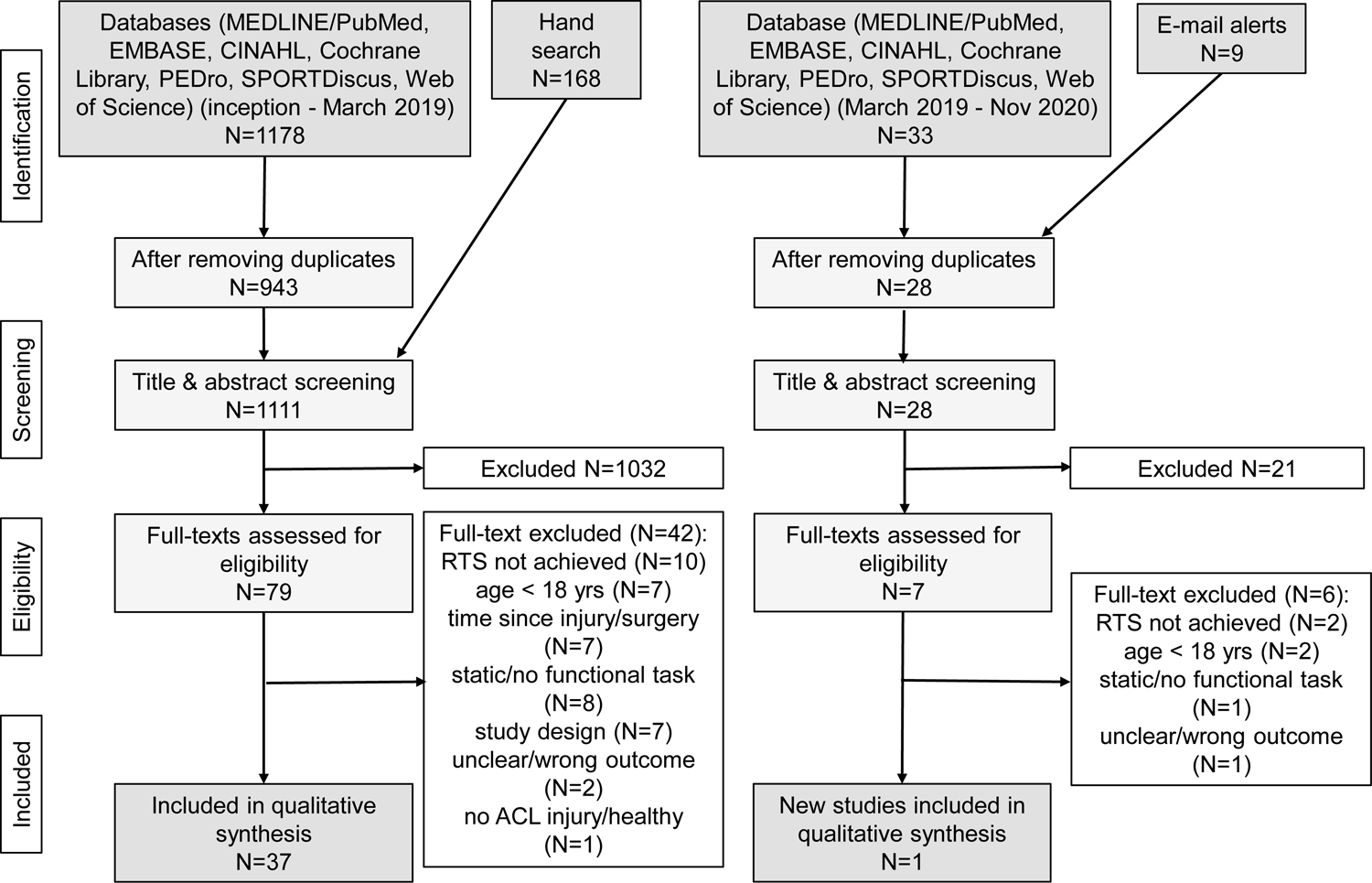
Flowchart of literature search according to guidelines of PRISMA (27) Legends: PEDro = Physiotherapy Evidence Database; PRISMA = Preferred Reporting of Items for Systematic Reviews and Meta-Analyses

### Risk of bias assessment

Risk of bias of half (19 studies, 50.0%) of the included studies was medium (13, 33–50), six (15.8%) showed high methodological quality (51–56) and 13 studies (34.2%) were of low quality (57–69) (Table 2). The main reasons for a medium to low methodological quality were due to an unclear description of participants and prior interventions, confounding factors, and incompletely reported results. Table 2 provides details about the risk of bias assessment for each included study.

**Table 2:**
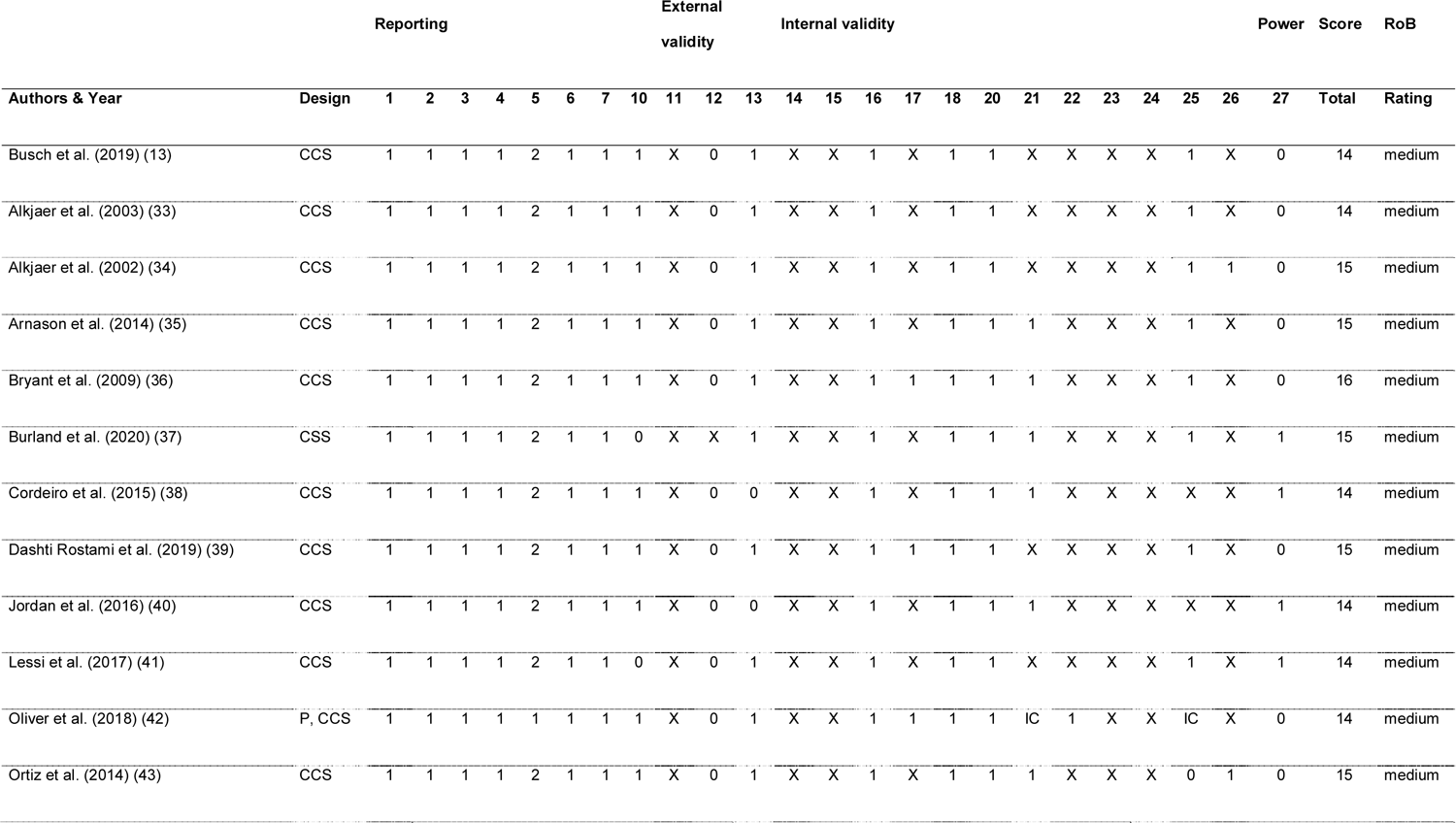
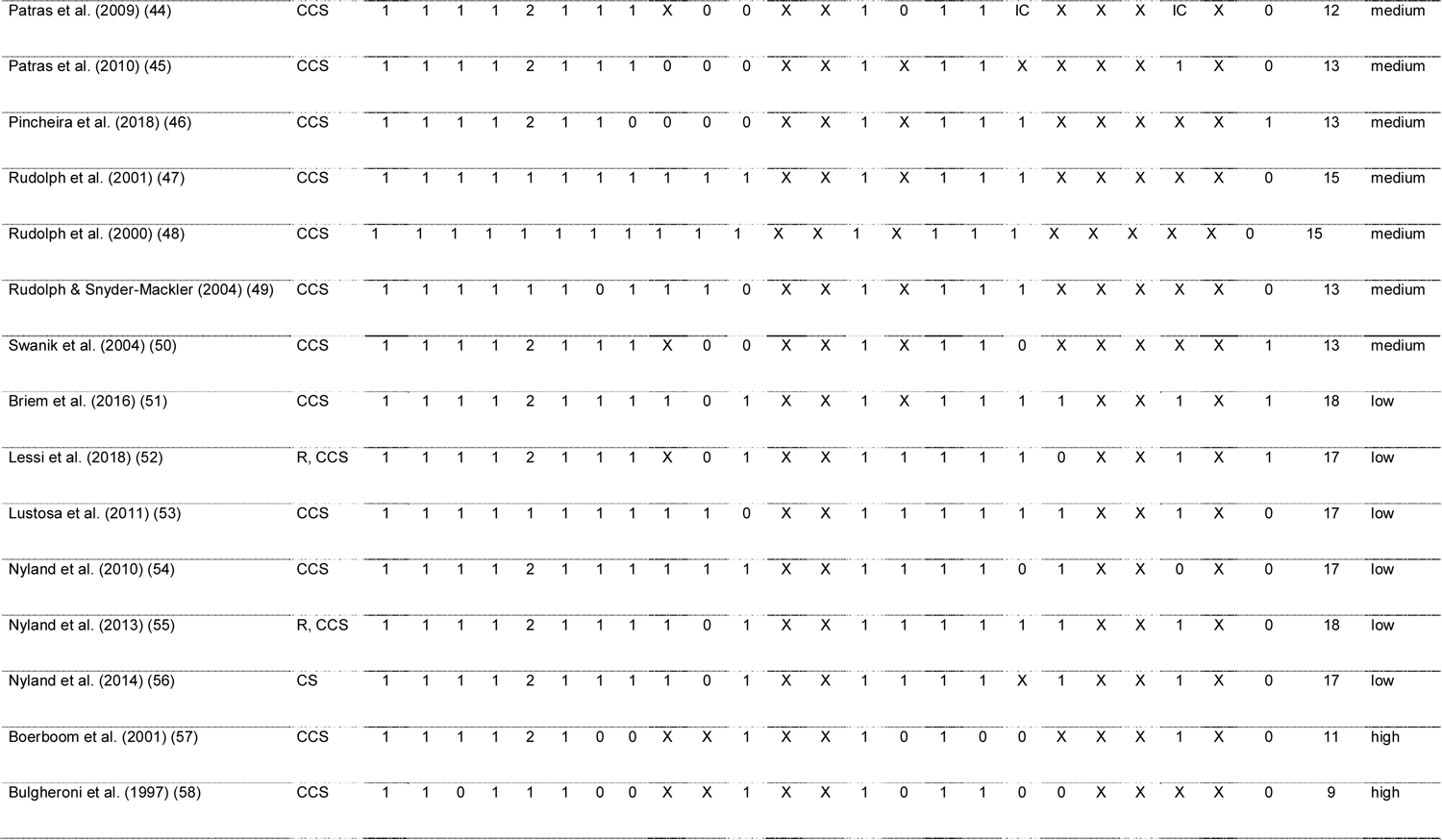
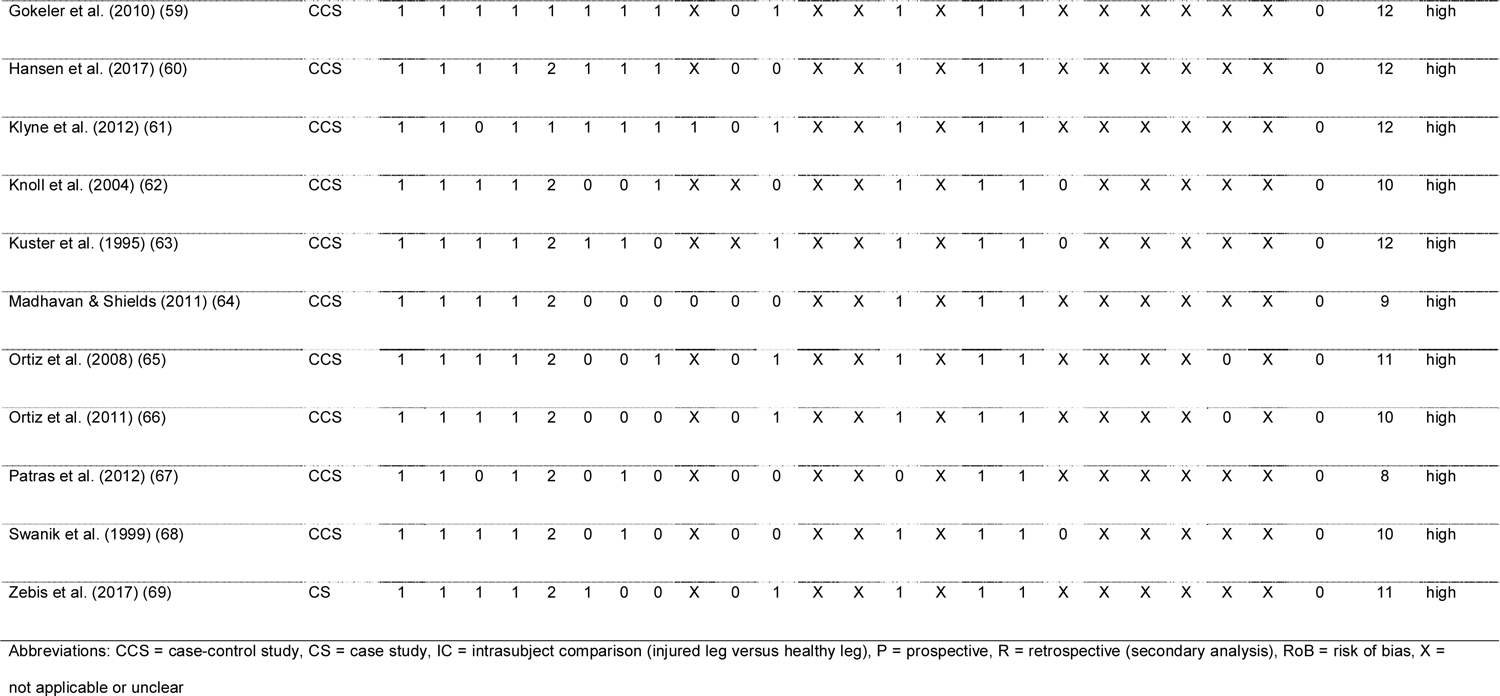
Risk of Bias assessment with modified Downs & Black checklist (29, 31, 32)

### Characteristics of included studies

*Study design:* All included studies were case-control studies, except two which where case series (56) or a single-case study (69). Two reported a retrospective or secondary data analysis (52, 55) or provided a subgroup analysis from a larger trial (45, 47–49, 65–67) (Table 2). Thirty-five studies compared the ACL participants with at least one control group (other ACL treatment, e.g. surgical versus conservative, or healthy controls), the remaining three studies made a comparison between the injured and the non-injured leg of the participants (42, 44) or compared the pre-injury status with follow-up data from pre- and post-surgery (69).

*Participants:* The number of included, adult participants with ACL injury varied from N = 1 (69) to a maximum of N = 70 (62) with a wide range of described physical activity from “normal” (58), “regular” (64), “active in at least one sport” (61), TAS ≥ 3 (50), minimal 2h/week (33, 34) to athletes at level I sports including jumping, pivoting and hard cutting (42, 57, 59), elite soccer players (35, 38, 67, 69) or elite skiers (50). Some authors restricted study participation to either males (33, 34, 36, 39, 44–46, 50, 58, 60, 67) or females (50, 51, 64–66, 68, 69), others measured females and males (13, 35, 37, 40, 41, 47–49, 52, 54–57, 59, 61, 62). Three studies did not provide any data about the gender of their participants (42, 53, 63). More patient characteristics of included studies can be found in Table 3.

**Table 3:** Participants’ characteristics of included studies

| Authors & Year | Number of participants (age, sex, group-specific inclusion criteria) | Diagnosis & treatment (only ACL) | Level of activity or sports (RTA, RTS, RTP) | Intervention Group | Control Group 1 (ACL patients) | Control Group 2 (healthy people) | significant difference between groups? |
| --- | --- | --- | --- | --- | --- | --- | --- |
| Busch et al. (2019) (13) | N = 20; N = 10 ACLR (age: 26 ± 10 yrs; height: 175 ± 6 cm; mass: 75 ± 14 kg) and N = 10 healthy matched controls (age: 31 ± 7 yrs; height: 175 ± 7 cm; mass: 68 ± 10 kg) | N = 10 ACLR (13.2 ± 2 months since repair), quadriceps tendon graft by same surgeon, some with additional injuries which needed surgery | TAS mind. 4 | N = 10 ACLR participants; age 26 ± 10 yrs, height 175 ± 7 cm, weight 75 ± 14kg, 3 females & 7 males, TAS 7 ± 2 | n.a. | N = 10 healthy participants without prior injury of the knee, age 31 ± 7 yrs, height 175 ± 8 cm, weight 68 ± 10kg, 3 females & 7 males, TAS 6 ± 1; matched according to age, height, weight, gender, (sports) activity level and leg dominance | no |
| Alkjaer et al. (2003) (33) | N = 29; N = 19, all male, complete chronic (post-injury time 6 months or more) ACLD and N = 10 healthy males as controls for EMG | complete chronic ACLD, min. 6 months of rehab program after injury, ACL injury clinically diagnosed by experienced orthopedic surgeons with Lachman, Anterior Drawer and Pivot-Shift Tests; TLS scores applied to separate ACLD-participants in copers and non-copers | min. 2h/wk of physical activity | N=9; male copers; (mass: 76.7 (14.3)kg, height: 1.81 (0.06) m, age: 28.3 (6.1) yrs); mean TLS scores: 87.1 (5.8) and 6.1 (0.6) respectively; mean time after injury: 39.1 (42.3) (range 6.0–120.0) months | N=10; male non-copers; mass: 80.4 kg (SD 6.7); height: 1.79 m (SD 0.05), age: 31.7 yrs (SD 5.9); mean TLS scores: 74.0 (SD 7.1) and 3.8 (SD 0.6), respectively; mean time after injury: 55.0 months (SD 42.7) (range 6.0–144.0) | N=10; male healthy; mass: 77.5 kg (SD 7.9), height: 1.82 m (SD 0.05), age: 31.0 yrs (SD 2.8) | no |
| Alkjaer et al. (2002) (34) | N = 23 all male; N = 17 males with complete ACLD, N = 6 healthy controls | complete ACLD, min. 6months of rehab program after injury; TLS scores applied to separate ACLD-participants in copers and non-copers | min. 2h/wk | N=8; male copers; weight: 76.6 kg (SD 14.8); height: 1.81 m (SD 0.06), age: 26.0 yrs (SD 4.0); mean TLS scores: 85.5 (SD 5.3) and 6.25 (SD 0.5), respectively, mean time after injury: 34.0 months (SD 39.2) (range 6.0–120.0) | N=9; male non-copers; weight: 80.6 kg (SD 7.1); height: 1.79 m (SD 0.06), age: 31.2 yrs (SD 6.0); mean TLS scores: 74.0 (SD 7.1) and 3.8 (SD 0.6), respectively; mean time after injury: 51.8 months (SD 44.0) (range 6.0–144.0) | N=6; male healthy; weight: 73.8 kg (SD 7.9), height: 1.81 m (SD 0.05), age: 31.0 yrs (SD 1) | no |
| Arnason et al. (2014) (35) | N = 36; N = 18, female and male soccer players with ACLR (post-injury time 1 – 6 yrs) and N = 18 healthy female and male soccer players from the same team (men's and women's top league in Iceland), matched for gender, height, body mass and "involved" side designation, as controls | ACLR: successful return to full participation in soccer; no muscle strain injury in knee flexors in past 3 months, no orthopedic condition excluding from soccer | full participation in soccer (Icelandic top leagues) | N=18 ACLR participants in total; N = 8 males, N = 10 females; all participants mean mass: 69.2 (11.8) kg, height: 1.73 (0.09)m, age: 23.7 (3.6) yrs; mean BMI 23.0 (2.4) kg/m2; left/right dominance 2/16; involved/uninvolved is the dominant leg 8/10; time since injury 1 – 6 yrs | n.a. | N=18 healthy participants; N = 8 males, N = 10 females; all participants mean mass: 68.6 (11.2) kg, height: 1.73 (0.08) m, age: 20.5 (3.7) yrs; mean BMI 22.7 (2.0) kg/m2 | no |
| Bryant et al. (2009) (36) | N = 59; N = 10 males with ACLD (18–35 yrs); N= 27 matched males with ACLR (14 with patella tendon graft, 13 with combined ST and gracilis graft); N = 22 matched-controls | Cincinnati Knee Rating System (0 – 100 points); ACLD: full ROM, neg. Lachman, neg. Pivot-Shift; confirmed isolated ACL rupture (arthroscopic) min. 1yr before testing; same orthopedic surgeon for all ACLR | n.m., but hopping required | N = 10 male with ACLD (18–35 yrs) | N= 27 matched males with ACLR (14 with patella tendon graft, 13 with combined ST and gracilis graft) | N = 22 matched (age, activity level, anthropometrics), healthy controls no history of trauma or disease in either knee & no evidence of abnormality on clinical examination | no |
| Burland et al. (2020) (37) | N = 36; N = 16 females ACLR, N = 10 males ACLR, N = 8 healthy controls (N = 4 females, N = 4 males) | unilateral ACLR, 21 subjects with BPTB graft, 5 with hamstrings graft; enrolled in physician-directed rehabilitation program, able to do single limb forward hop | Not mentioned as criteria for in-/exclusion; minimum TAS of 5 preinjury/ after surgery | N = 26 with ACLR, age 20.2 ± 2.7 yrs, mean time since injury 2.2 ± 2.7 yrs, TAS preinjury median 8.0 (range 5.0–10.0), after surgery 7.0 (range 5.0–9.0) points, cleared for unrestricted RTS | n.a. | N = 8 healthy controls, age 23.3 ± 1.8 yrs, TAS median 9.0 (range 5.0–10.0) points | no, except age |
| Cordeiro et al. (2015) (38) | N = 17 males; N = 8 with ACLR and N = 9 healthy controls | ACLR: mind. 6 months post-surgery on dominant leg, bone-tendon-bone arthroscopy, no problems at end of physiotherapy phase | soccer, professional level | N = 8 N = 8 professional male soccer players (age = 24.6 ± 3.5 yrs, height = 1.83 ± 0.06 m, mass = 77.3 ± 7 kg) with ACLR mind. 6 months since surgery | n.a. | N = 9 healthy controls; professional male soccer players (age = 24.0 ± 3.5 yrs, height = 1.76 ± 0.05 m, mass = 72.9 ± 3.5 kg), no knee or leg injuries or previous ACL surgeries | no |
| Dashti Rostami et al. (2019) (39) | N = 36; N = 12 ACLD, N = 12 ACLR; N = 12 healthy controls; all male athletes | for patients: primary unilateral ACL injury | athletes, regular sports participation, ACLD = copers | N = 12 males, 18 to 36 months post-ACLR | N = 12 males, 18 to 36 months after ACL rupture (= ACLD, copers); grade 2 or 3 rupture including the following definition of copers: athletes with ACLD for at least 18 months, no symptoms of knee instability during regular sports participation | N = 12 healthy males, matched controls; no knee injury, no knee pain | no |
| Jordan et al. (2016) (40) | N = 22; N = 11 ACLR, N = 11 control; elite skiing athletes from Canada's national alpine skiing and skier cross team | ACLR: primary ACL injury, at least 12 months post-surgery, actively competing athletes at the Federation International de Ski World Cup level with full medical clearance to compete | elite ski racers, TAS 10, competing at international level | N = 11 actively competing ACLR skiers (females, n = 5: age = 23.6 ± 1.8 yrs, mass = 61.0 ± 5.3 kg; males, n = 6: age = 26.5 ± 5.8 yrs, mass = 84.4 ± 9.0 kg; 7 subjects with ST autograft, 1 with BPTB autograft, 3 with cadaver allograft | n.a. | N = 11 matched controls with no history of ACL injury (females, n = 5: age = 21.8 ± 3.2 yrs, mass = 63.7 ± 4.6 kg; males, n = 6: age = 23.3 ± 3.3 yr, mass = 84.7 ± 5.1 kg; active competitors at the international level defined as participation in the Federation International de Ski World Cup circuit | n.m. |
| Lessi et al. (2017) (41) | N = 40; N = 20 with ACLR, N = 20 healthy controls | ACLR: non-contact ACL injury, unilateral reconstruction of the ACL with no prior history of a contralateral ACL injury, no recent history of an ankle, hip, spine, or contralateral knee injury in the past 12 months; rehabilitation completed, cleared to RTS by both their physician and physical therapist | recreational sports, meaning aerobic or athletic activity at least 3x/wk | N = 20 with ACLR, 13 males, 7 females, at least 12 months post-surgery, 13 with hamstring ipsilateral autografts, 7 with BPTB ipsilateral autograft), | n.a. | N = 20 healthy controls, 13 males, 7 females, no history of any dysfunction or previous joint trauma, no prior history of ACL injury or injury of lower extremity in last 12 months; were matched by age, sex, weight, and current sporting activity type | no |
| Oliver et al. (2018) (42) | N = 25 ACLD, mean age $22 \pm 4.61$ yrs, mean mass $71.18 \pm 10.57$ kg, mean height was $177.55 \pm 9.69$ cm; N = 18 males (72%); N = 2 lost to follow-up due to personal issues, all remaining 23 patients concluded the study (pre-surgery, 4 and 6 months post-surgery for questionnaires, at 6 months for jumps) | complete ACL tear was based on clinical symptoms, on positive Lachman's and pivot shift tests, and was confirmed by magnetic resonance imaging; reconstruction 2–3 months after the injury by same surgeon using BTB-technique | more than 200 h of sports activity per year, including jumping, pivoting and twisting actions | injured knee | non-injured knee | n.a. | n.a. |
| Ortiz et al. (2014) (43) | N = 31 females; N = 15 ACLR, N = 16 healthy females | ACLR: same orthopedic surgeon, same rehabilitation protocol, N = 13 were injured while participating in competitive volleyball at the collegiate or professional level; at least 12 months post-surgery, full RTS allowed (without restrictions) to pre-injury level | sports-specific physical activities as described by the Activity Rating Scale, scores from 12 to 16, consistent with activities such as running, cutting, decelerating, and pivoting more than 2x/wk = high level of participation | N = 15 ACLR with SG graft, age range 21 – 35 yrs (height: $167.71 \pm 9.0$ cm, body mass: $67.68 \pm 11.66$ kg), time since surgery was between 12 months and 5 yrs, full RTS allowed (pre-injury level); N = 1 drop-out due to inability to perform tasks | n.a. | N = 16 healthy females, participating in volleyball, basketball, and soccer at the collegiate or intramural sports level, age range 21 – 35 yrs, height: $160.50 \pm 5.17$ cm, body mass: $59.35 \pm 10.37$ kg | no for age and activity, height and weight |
| Patras et al. (2009) (44) | N = 9 males with ACLR | ACLR: unilateral ACL tear confirmed by MRI and arthroscopy, BPTB graft within 6 months after injury, same rehabilitation protocol, RTS permitted 6 months post-surgery | athletes, amateur soccer players, at least TAS 7 | N = 9 males with ACLR, mean age: $27.7 \pm 3.5$ yrs, mean weight: $79.5 \pm 7.3$ kg, mean height: $178 \pm 5.9$ cm, mean time since surgery: $19.2 \pm 5.7$ months, median Lysholm score: 95 (range 94–96), TAS: 8 (range 7–9), resumed their sports activities | n.a. | n.a., non-injured side respectively | n.a. |
| Patras et al. (2010) (45) | N = 28 males; N = 14 ACLR, N = 14 healthy controls | ACLR: unilateral ACL tear confirmed by MRI and arthroscopy, BPTB graft, performed within 6 months after injury, same surgeon, same rehabilitation, RTS permitted after 6 months post-surgery | amateur soccer players | N = 14 males with ACLR, mean age: $24.8 \pm 5.3$ yrs; mean height: $177 \pm 5.3$ cm, mean weight $77.3 \pm 7.5$ kg, time since surgery: mean $18.5 \pm 4.3$ months, pre-injury level of sports participation, median Lysholm score 95 (range 94–100) and TAS 8 (range 7–9) | n.a. | N = 14 healthy males, mean age: $21.7 \pm 4.4$ yrs; mean height: $180 \pm 9.0$ cm, mean weight $72.2 \pm 8.3$ kg, never suffered of any kind of orthopedic or neurological condition; left leg = control leg | n.m. |
| Pincheira et al. (2018) (46) | N = 50 male soccer players; N = 25 with unilateral ACLR, N = 25 uninjured controls | ACLR: unilateral ACLR with ST-gracilis graft, same surgical team, at least 6 months post surgery; non-contact mechanism during soccer match on the dominant limb | amateur soccer players, playing at least 2x/wk | N = 25 males with ACLR, age $28.36 \pm 7.87$ yrs; weight $77.56 \pm 6.35$ kg, height $169 \pm 7$ cm, time after surgery $9 \pm 3$ months, time between ACL injury and surgery $3.4 \pm 1$ months; at time of measurements cleared for full RTS | n.a. | N = 25 healthy males, age $24.16 \pm 2.67$ yrs; weight $78.16 \pm 5.46$ kg, height $172 \pm 5$ cm; without injury or surgery on lower limb | no |
| Rudolph et al. (2001) (47) | one component of a larger study; N = 31; N = 10 healthy controls, N = 11 ACLD copers, N = 10 ACLD non-copers | ACLD: full range of motion in both knees, no visible or palpable knee effusion, no symptoms of locking, an uninvolved, healthy knee | athletes, regular activity in level I sports (involving jumping, pivoting, and hard cutting) and level II sports (involving lateral motions) before injury | N = 11 ACLD copers (2 females, 9 males), ages 22–43 yrs, mean 30.7, high-level athletes with ACLD for at least 1 year (confirmed by MRI, any knee instability during regular participation in level I and II sports, no more than one episode of giving way, even during sports, since injury | N = 10 noncopers ACLD (4 females, 6 males), ages 16–43 yrs, mean 28.1; more than one episode of giving way since injury, instability during ADL, not returned to sports | N = 10 uninjured individuals, matched by age and activity level to the coper subjects (2 females, 8 men), ages 23–41 yrs, mean 32.2) | no (age and joint laxity) |
| Rudolph et al. (2000) (48) | one component of a larger study; N = 31; N = 10 healthy controls, N = 11 ACLD copers, N = 10 ACLD non-copers | ACLD: full range of motion in both knees, no visible or palpable knee effusion, no symptoms of locking, an uninvolved, healthy knee | athletes, regular activity in level I sports (involving jumping, pivoting, and hard cutting) and level II sports (involving lateral motions) before injury | N = 11 ACLD copers (2 females, 9 males), ages 22–43 yrs, mean 30.7, high-level athletes with ACLD for at least 1 year (confirmed by MRI, any knee instability during regular participation in level I and II sports, no more than one episode of giving way, even during sports, since injury | N = 10 noncopers ACLD (4 females, 6 males), ages 16–43 yrs, mean 28.1; more than one episode of giving way since injury, instability during ADL, not returned to sports | N = 10 uninjured individuals, matched by age and activity level to the copers (2 females, 8 men), ages 23–41 yrs, mean 32.2) | n.m. |
| Rudolph & Snyder-Mackler (2004) (49) | one component of a larger study; N = 31; N = 10 healthy controls, N = 11 ACLD copers, N = 10 ACLD non-copers | ACLD: full range of motion in both knees, no visible or palpable knee effusion, no symptoms of locking, an uninvolved, healthy knee | athletes, regular activity in level I sports (involving jumping, pivoting, and hard cutting) and level II sports (involving lateral motions) before injury | N = 11 ACLD copers (2 females, 9 males), ages 22–43 yrs, mean 30.7, high-level athletes with ACLD for at least 1 year (confirmed by MRI, any knee instability during regular participation in level I and II sports, no more than one episode of giving way, even during sports, since injury | N = 10 noncopers ACLD (4 females, 6 males), ages 16–43 yrs, mean 28.1; more than one episode of giving way since injury, instability during ADL, not returned to sports | N = 10 uninjured individuals, matched by sex, age and activity level to the copers (2 females, 8 men), ages 23–41 yrs, mean 32.2) | no (age and leg length) |
| Swanik et al. (2004) (50) | N = 29; N = 12 female ACLD, N = 17 female controls | complete unilateral ACL tear, at least 1 year after injury, mechanical instability (positive Lachman & Pivot-Shift tests), rehabilitation program completed, no ACL surgery | minimum TAS of 3 | N = 12 females with ACLD, age $25.2 \pm 7.3$ yrs, mean time since injury $33.6 \pm 5.2$ months, TAS $5.4 \pm 1.83$ points | n.a. | N = 17 healthy females, age $22.7 \pm 4.0$ yrs, TAS $5.41 \pm 1.5$ points | n.m. |
| Briem et al. (2016) (51) | N = 36; N = 18, female players with ACLR (post-injury time 1 – 6 yrs) and N = 18 healthy female players from the same team (from Icelandic women's top league in handball, football, basketball), matched for gender, height, body mass and "involved" side designation, as controls | no information about diagnosis or treatment; exclusion criteria: current musculoskeletal injury, lower limb muscle strain within 3 previous months, not being able to do single-limb hops | ACLR: successful return to competition with their teams; healthy: full participation in soccer (Icelandic top leagues) | N = 18 females, ACLR, recruited via advertisement from teams competing in the tip leagues in three sports team handball (n = 5), basketball (n = 4), and football (n = 9)]. In 12 instances, the surgical limb was the individual's dominant one. Characteristics: mean mass: 67.2 (7.8) kg, height: 1.714 (0.05) m, age: 22.7 (3.5) yrs; mean BMI 22.8 (2.4) kg/m<sup>2</sup>; involved/uninvolved is the dominant leg 12/18; time since injury 1 – 6 yrs | n.a. | N = 18 healthy females recruited from the same teams, matched for age, height, weight. Characteristics mean mass: 66.3 (7.1) kg, height: 1.708 (0.05) m, age: 21.5 (2.7) yrs; mean BMI 22.7 (2.2) kg/m<sup>2</sup> | no |
| Lessi et al. (2018) (52) | N = 14 ACLR (7 males, 7 females) from study of Lessi et al. (2017) (41) | non-contact ACL injury; unilateral ACLR with autologous ipsilateral graft at least 12 months before recruitment; undergone a rehabilitation program; returned to sports participation; no contralateral ACL injury | recreational sports | N = 7 males ACLR, age 23.90 ± 2.80 yrs, height 1.80 ± 0.1m, body mass 83.3 ± 7.8 kg, 3 with BPTB graft, 4 with flexor tendons grafts | N = 7 females ACLR, age 24.7 ± 5.3 yrs, height 1.63 ± 0.1m, body mass 65.9 ± 9.0 kg, 2 with BPTB graft, 5 with flexor tendons grafts | n.a. | no, except men were taller than women (P < 0.001) and performed a higher number of sets of the protocol before becoming fatigued their reconstruct ed limb (P = 0.006). |
| Lustosa et al. (2011) (53) | N = 25 ACLR; N = 15 with Cincinnati Knee Rating System (CKRS) > 90 points (full RTS), N = 10 with CKRS < 85 points (limited RTS) | at least 2 yrs post-surgery, same rehabilitation program which allowed full RTS activities 7 months post-surgery | full RTS allowed, not further specified | N = 10 ACLR with CKRS 77.30 ± 6.14 points, age 33.4 ± 7.53 yrs, time between injury and surgery 52.20 ± 31.33 months, 3 with associated meniscal injuries, 7 without | N = 15 ACLR with CKRS 96.87 ± 2.75 points, age 34.5 ± 8.85 yrs, time between injury and surgery 67.3 ± 28.5 months, 3 with associated meniscal injuries, 12 without | n.a. | no |
| Nyland et al. (2010) (54) | N = 70 ACLR; N = 35 males; N = 35 females, 5.3 ± 3 yrs post-surgery | minimum of 2 yrs since unilateral primary ACL reconstruction with allografts performed by same surgeon, standard rehabilitation program with sufficient adherence | met or exceeded standard accepted return-to-sports activity goals of a minimum 85% bilateral equivalence with single-leg hop-for-distance testing and 60°/s isokinetic peak knee extensor and flexor torque testing | N = 35 males with ACLR, age n.m., height 180.3 ± 6.9cm, weight 88.9 ± 13.3kg, time after surgery 5.6 ± 3.2 yrs | N = 35 females with ACLR, age n.m., height 166.6 ± 7.1cm, weight 68.2 ± 18.9kg, time after surgery 5.1 ± 2.6 yrs | n.a. | n.m. |
| Nyland et al. (2013) (55) | N = 70 ACLR; 35 male and 35 females, 5.3 ± 3 yrs after surgery; secondary analysis of Nyland et al., 2010 (54) | minimum of 2 yrs since unilateral primary ACL reconstruction with allografts performed by same surgeon, standard rehabilitation program with sufficient adherence | met or exceeded standard accepted return-to-sports activity goals of a minimum 85% bilateral equivalence with single-leg hop-for-distance testing and 60°/s isokinetic peak knee extensor and flexor torque testing | N = 24 ACLR well-trained/frequently sporting, 50% males, age at surgery 29.8 ± 11.4 yrs, height 172.5 ± 8.6 cm, weight 77.1 ± 18.2 kg, time post-surgery 5.7 ± 2.8 yrs, IKDC 87.3 ± 11.5 | N = 26 ACLR only sporting sometimes, 50% males, age at surgery 33.1 ± 13.5 yrs, height 171.7 ± 9.7 cm, weight 79.4 ± 23.2 kg, time post-surgery 5.4 ± 3.1 yrs, IKDC 87.3 ± 11.5 | no healthy control group, but N = 20 ACLR highly competitive subjects, 50% males, age at surgery 26.5 ± 9.4 yrs, height 176.5 ± 9.4 cm, weight 76.8 ± 13.9 kg, time post-surgery 4.6 ± 3.0 yrs, IKDC 91.0 ± 9.4 | no |
| Nyland et al. (2014) (56) | N = 65 ACLR; 32 male and 33 females, 5.2 ± 2.9 yrs after surgery; Subject group assignments were made based on how they responded to the following question: "Compared to prior to your knee injury how capable are you now in performing sports activities", very capable (group 1 → see field for healthy controls), capable (group 2), or not capable (group 3) | minimum of 2 yrs since unilateral primary ACL reconstruction with allografts performed by same surgeon, standard rehabilitation program with sufficient adherence | met or exceeded standard accepted return-to-sports activity goals of a minimum 85% bilateral equivalence with single-leg hop-for-distance testing and 60°/s isokinetic peak knee extensor and flexor torque testing | N = 23 "capable = group 2", 52.2% males, age at surgery 29.3 [95% CI: 24.1, 34.4] yrs, height 172.8 [168.4, 177.3] cm, weight 76.8 [68.3, 85.2] kg, time post-surgery 5.4 [4.2, 6.6] yrs, IKDC 87.2 [82.1, 92.4] | N = 22 "not capable = group 3", 45.5% males, age at surgery 33.6 [95% CI: 26.4, 39.1] yrs, height 172.1 [167.1, 177.1] cm, weight 79.7 [68.0, 91.3] kg, time post-surgery 5.2 [3.8, 6.5] yrs, IKDC 78.6 [71.7, 85.5] | no healthy control group, but N = 20 "very capable = group 1", 50% males, age at surgery 26.5 [95% CI: 21.9, 31.8] yrs, height 176.5 [170.4, 180.1] cm, weight 76.8 [67.4, 80.3] kg, time post-surgery 4.6 [2.8, 6.2] yrs, IKDC 91.0 [84.1, 94.6] | no |
| Boerboom et al. (2001) (57) | N = 20; N = 10 ACLD (5 copers, 5 noncopers), N = 10 controls | ACLD: ACL rupture confirmed by physical examination and arthroscopy, conservative treatment | before injury: all ACLD participants at level I (of the IKDC score), after injury: level I (all copers), level II and III (noncopers) | N = 5 copers (all males) with ACLD, median age 32 yrs, range 21–46, median time between primary injury and gait analysis: 39 months (13–67), acting at same level of sports and daily activities (level I) as before the injury | N = 5 noncopers (3 males, 2 females) with ACLD, with functional instability, median age 27 yrs, range 23–35, median time between injury and gait analysis: 22 months (16–87), acting at lower level (4 at level III, 1 at level II) | N = 10 healthy males, without a history of knee injury, median age was 22 yrs (range 18–24) | no in patient groups (age, time between injury and gait analysis), in comparison with healthy controls: n.m. |
| Bulgheroni et al. (1997) (58) | N = 30 all males; N = 15 with ACLR, N = 10 with ACLD, N = 5 healthy controls | ACLR: BPTB graft | normal activity | N = 15 males with ACLR, age 25 ± 3 yrs, time after reconstruction: 17 ± 5 months, normal activity | N = 10 males with ACLD, age 27 ± 6 yrs, mean time after injury: 20.4 months after injury (range 8–48 months), knee instability | N = 5 males, healthy controls, age 28 ± 3 yrs, no history of musculoskeletal pathology | n.m. |
| Gokeler et al. (2010) (59) | N = 20; N = 9 ACLR patients, N = 11 healthy controls | ACLR: six months after surgery, isolated ACL lesion, no major meniscal or cartilage lesion, normal limb alignment, no relevant previous surgery at any other joint of the limbs, same rehab program at same institution, unrestricted RTS allowed after 9 months post-surgery | level I-II athletes | N = 9 ACLR patients (6 males, 3 females), mean age 28.4 ± 9.7 yrs, 27 ± 1.5 wk postoperatively (BPTB technique, same surgeon), | n.a. | N = 11 healthy subjects (8 males, 3 females), level I-II athletes, | n.m. |
| Hansen et al. (2017) (60) | N = 37; N = 18 male patients, N = 19 healthy participants | ACLR: discharged from rehabilitation facility, | ready to return to on-fields sports specific activity | N = 18 male ACLR at the end of their rehabilitation and allowed to running, 7 ± 2 months post-surgery; N = 8 with a BPTB graft, age 27 ± 7.69 yrs, weight 80.40 ± 9.44 kg, height: 178.49 ± 7.29 cm; N = 10 with a hamstring graft, age 26 ± 3.84 yrs, weight 74.16 ± 7.19 kg, height 176.89 ± 5.6 cm | n.a. | N = 19 injury-free male controls, age 35.4 ± 7.8 yrs, weight 77.6 ± 8.4 kg, height 179.1 ± 5.6 cm | n.m. |
| Klyne et al. (2012) (61) | N = 26; N = 15 ACLD, N = 11 healthy controls | ACLD: chronic, unilateral ACL rupture demonstrated with a positive pivot shift and confirmed by orthopedic surgeon, plus a history of subjective stability and a right skill preference in the lower limb, without previous ACL surgery | active in at least one sport | N = 15 ACLD, 10 males and 5 females, 28 ± 7 yrs, average time since injury of 34 months (±17 months), sustained injury while playing sport | n.a. | N = 11 healthy controls, 9 males, 2 females (29 ± 8 yrs), active in at least one sport, no other musculoskeletal problems, right skill preferred in their lower limb, matched for age and activity level | n.m. |
| Knoll et al. (2004) (62) | N = 76; N = 25 ACLR (pre- and postsurgery), N = 51 healthy controls | no previous injury, no meniscal damage, BPTB graft, rehabilitation program | non-professional athletes pursuing some sports two to three times/wk | N = 25 with ACLD (before surgery, later ACLR), 18 males, 7 females; first subgroup: 9 male with acute ACLD (mean age: 29.86 ± 6.52 yrs, mean height 1.77 ± 0.8 meters, mean mass 81.40 kg ± 9.06 kg); second subgroup: 9 males with chronic ACLD (mean age: 39.70 ± 2.1 yrs mean height: 1.70 ± 0.21 meters, mean mass: 88.1 ± 20.2 kg) and 7 females with chronic ACLD (mean age: 30.31 ± 9.48 yrs, mean height: 1.64 ± 0.32 meters, mean mass: 62.0 ± 8.4 kg). The chronic ACLD group was examined an average of<br>28.2 months after injury<br>(ranging from 24 to 52<br>months), but before<br>surgery | same population of ACLD, but after surgery → ACLR, measured at wk 6, and 4, 8, and 12 months post-surgery | N = 51 healthy controls, 31 males, 20 females, mean age: 31.70 ± 4.1 yrs, mean height: 1.71 ± 0.12 meters, mean mass 72.1 ± 25.2 kg, no pathology that would affect gait, unfamiliar with treadmill walking | n.m. |
| Kuster et al.<br>(1995) (63) | N = 33; N = 21 with<br>ACLD, N = 12 healthy<br>controls | ACLD: arthroscopically<br>confirmed complete<br>ACL ruptures at least 1<br>year previously | ACLD: TAS 6-10 (mean 8.2)<br>before injury<br>and 3-9 (mean<br>5.3) after injury;<br>controls: TAS<br>range 4-8<br>(mean 6.1) | N = 19 with 21 ACLD,<br>mean age: 28.2 yrs<br>(range 19-42), mean<br>height: 174.1cm (156-187.6), mean weight:<br>77.9kg (50-112), mean<br>time since injury: 45<br>months (range of 12-108), mean Lysholm<br>score 82 (range 55-100), | n.a. | N = 12 healthy controls,<br>similar in height and weight,<br>mean height: 171.2 cm and<br>weight 70.8 kg, no lower limb<br>injury | unclear<br>(similar for<br>height and<br>weight) |
| Madhavan &<br>Shields (2011)<br>(64) | N = 24 females; N =<br>12 with ACLR, N = 12<br>healthy controls | complete reconstruction<br>of the ACL with BPTB<br>or HS autograft, ability<br>to climb stairs without<br>difficulty, full joint ROM,<br>SR-36, KOOS, IKDC | regular physical<br>activity, TAS | N = 12 females ACLR,<br>age $22.4 \pm 2.4$ yrs, mean<br>time from surgery $3.7 \pm$<br>$1.8$ yrs, weight $144.1 \pm$<br>$19$kg, height $164.5 \pm$<br>$5.28$cm, TAS (current) $7.1$<br>$\pm 2.4$ | n.a. | N = 12 healthy females, no<br>previous history of knee<br>pathology, age $24.1 \pm 3.2$ yrs,<br>weight $136.5 \pm 20.3$kg, height<br>$163.8 \pm 7.3$cm, TAS (current)<br>$6.9 \pm 2.1$; matched to age | no |
| Ortiz et al. (2008)<br>(65) | N = 28 females; N =<br>13 ACLR, N = 15<br>non-injured controls | not controlled for<br>graft/surgery or<br>rehabilitation protocol<br>(only similarities); at<br>least 1-year post<br>surgery, no multiple<br>surgeries on the same<br>knee | recreational<br>fitness activities<br>such as<br>jogging,<br>running, and<br>weightlifting,<br>none of the<br>participants<br>formed part of<br>any<br>intercollegiate,<br>varsity, or<br>competitive<br>sport team | N = 14 physically active<br>young women with ACLR<br>(age, $25.4 \pm 3.1$ yrs;<br>height, $167.5 \pm 5.9$ cm;<br>body mass, $63.2 \pm 6.7$ kg;<br>mean time after surgery<br>$7.2 \pm 4.2$ yrs (1–16 yrs<br>after reconstruction); N =<br>9 with BPTB graft, N = 3<br>with gracilis-ST-graft, N =<br>2 with Achilles tendon<br>graft; N = 1 excluded due<br>to inability to perform<br>tasks | n.a. | N = 15 healthy, noninjured<br>young women from<br>physiotherapy school (age,<br>$24.6 \pm 2.6$ yrs; height, $164.7 \pm$<br>$6.5$ cm; body mass, $58.4 \pm 8.9$<br>kg | n.m. |
| Ortiz et al. (2011)<br>(66) | N = 28 females; N =<br>13 ACLR, N = 15<br>non-injured controls<br>(same group as for<br>Ortiz et al., 2008 (65)) | not controlled for<br>graft/surgery or<br>rehabilitation protocol<br>(only similarities); at<br>least 1-year post<br>surgery, no multiple<br>surgeries on the same | recreational<br>fitness activities<br>such as<br>jogging,<br>running, and<br>weightlifting,<br>none of the<br>participants | N = 14 physically active<br>young women with ACLR<br>(age, $25.4 \pm 3.1$ yrs;<br>height, $167.5 \pm 5.9$ cm;<br>body mass, $63.2 \pm 6.7$ kg;<br>mean time after surgery<br>$7.2 \pm 4.2$ yrs (1–16 yrs<br>after reconstruction); N = | n.a. | N = 15 healthy, non-injured<br>young women from<br>physiotherapy school (age,<br>$24.6 \pm 2.6$ yrs; height, $164.7 \pm$<br>$6.5$ cm; body mass, $58.4 \pm 8.9$<br>kg | n.m. |
|  |  | knee | formed part of any intercollegiate, varsity, or competitive sport team | 9 with BPTB graft, N = 3 with gracilis-ST-graft, N = 2 with Achilles tendon graft; N = 1 excluded due to inability to perform tasks |  |  |  |
| Patras et al. (2012) (67) | N = 28 males; N = 14 ACLR and N = 14 healthy controls | ACLR: performed sub-acutely within 6 months after the injury from the same surgeon (range 1 to 4 months), unilateral ACL tear confirmed by MRI and arthroscopy; full RTS allowed 6 months post-surgery | competitive soccer players | N = 14 ACLR with BPTB autograft, age $24.8 \pm 5.3$ yrs, weight $77.3 \pm 7.5$ kg and height $177 \pm 5.3$ cm, mean time since surgery $18.5 \pm 4.3$ months; TAS 8 (range 7-9), Lysholm score 95 (range 94 – 100). | n.a. | N = 14 healthy male controls, age $21.7 \pm 4.4$ yrs, weight $72.2 \pm 8.3$ kg and height $180 \pm 9.0$ cm | n.m. |
| Swanik et al. (1999) (68) | N = 24 females, mean age = $29.4 \pm 10.4$ yrs; mean height = $168 \pm 10.7$ cm; mean weight = $61.2 \pm 6$ kg; N = 6 ACLD, N = 12 ACLR, N = 6 controls | complete unilateral ACL tear, ACLR: BPTB grafts, testing 6 – 30 months after surgery, rehabilitation program completed, attempt to previous level of activity | recreational activity at least for healthy controls, TAS of experimental groups $6.8 \pm 1.5$ points, Lysholm Knee Scoring Scale of experimental groups $92.9 \pm 5.4$ | N = 6 females with ACLD | N = 12 females with ACLR | N = 6 females, healthy controls, recreational activity, no previous history of knee pathology, dominant limb (leg to kick a ball with) | n.m. |
| Zebis et al. (2017) (69) | N = 1 female, age: 21 yrs | non-contact ACL injury (video-recorded) in the right knee during match play, ST-gracilis graft, standardized rehabilitation | elite soccer players | N = 1 female elite soccer player at high level with no previous history of ACL injury | n.a. | screening of elite soccer players pre-season | n.a. |

Legends: ACLD = anterior cruciate ligament deficiency (conservative/non-surgical treatment); ACLR = anterior cruciate ligament reconstruction/repair (surgery); BPTB = bone-patella-tendon-bone technique for ACLR; CI = confidence interval; IKDC = International Knee Documentation Committee; Level I: sports are described as jumping, pivoting and hard cutting sports; Level II sports: also involve lateral motion, but with less jumping or hard cutting than level I; n.a. = not applicable; n.m. = not mentioned; RTA = return to activity (return to participation); RTS = return to sports; RTP = return to performance; SD = standard deviation; ST = semitendinosus muscle; TAS = Tegner Activity Score; TLS = Tegner & Lysholm Score; TSK = Tampa Scale for Kinesiophobia; vs. = versus; wk = week; yrs = years

Details regarding methodological aspects of all included studies are presented in Table 4 below.

**Table 4.**
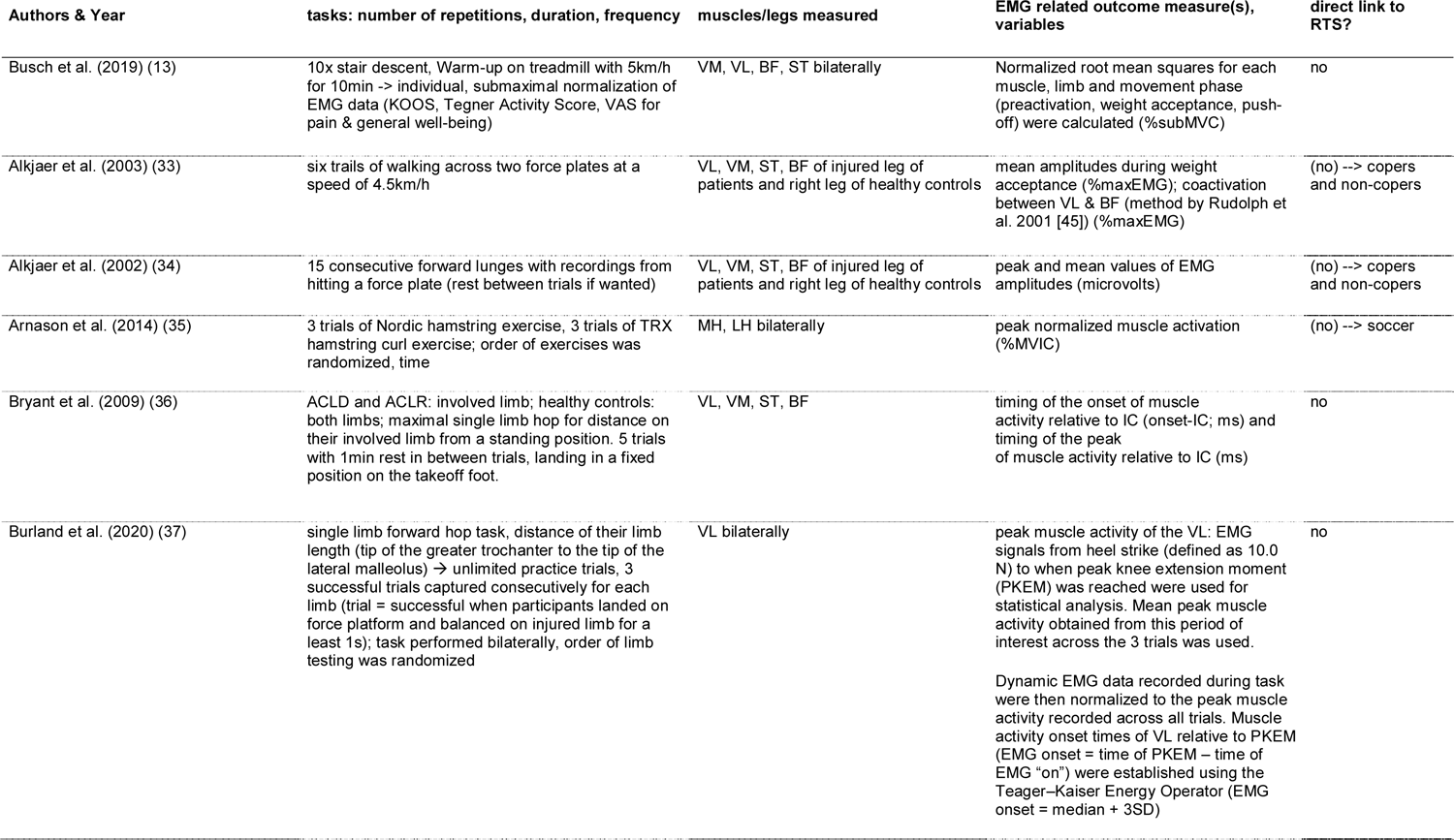
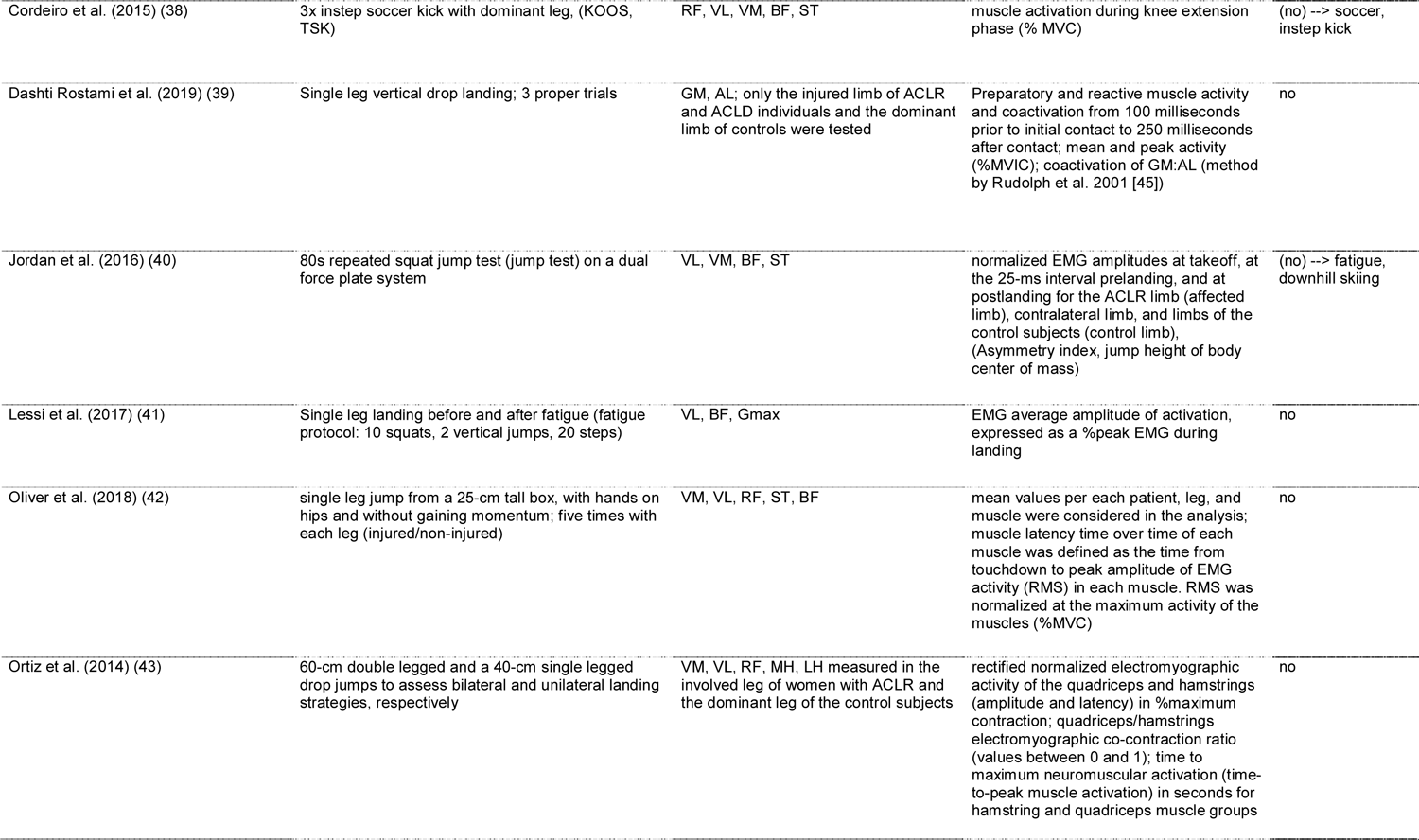
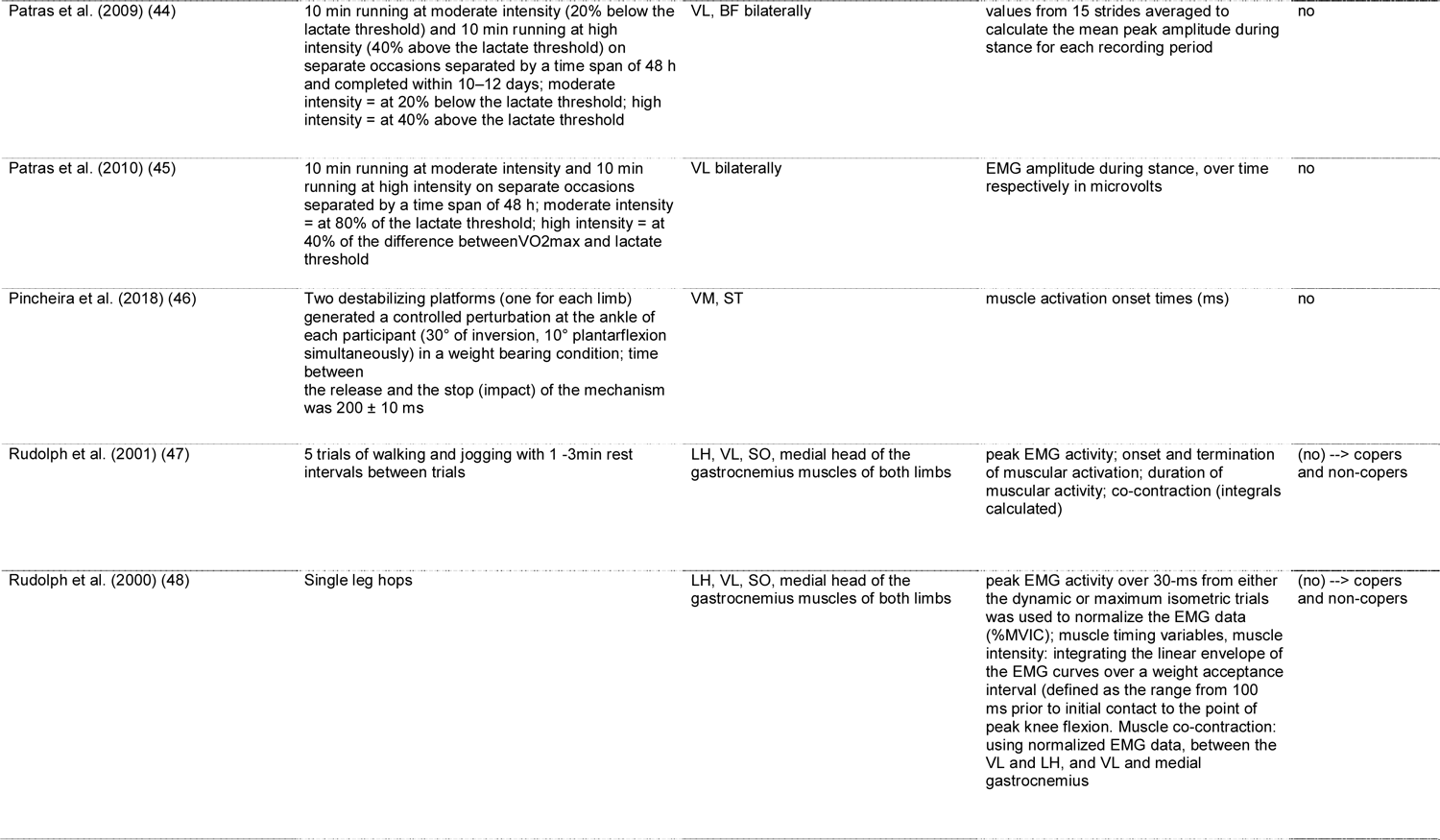
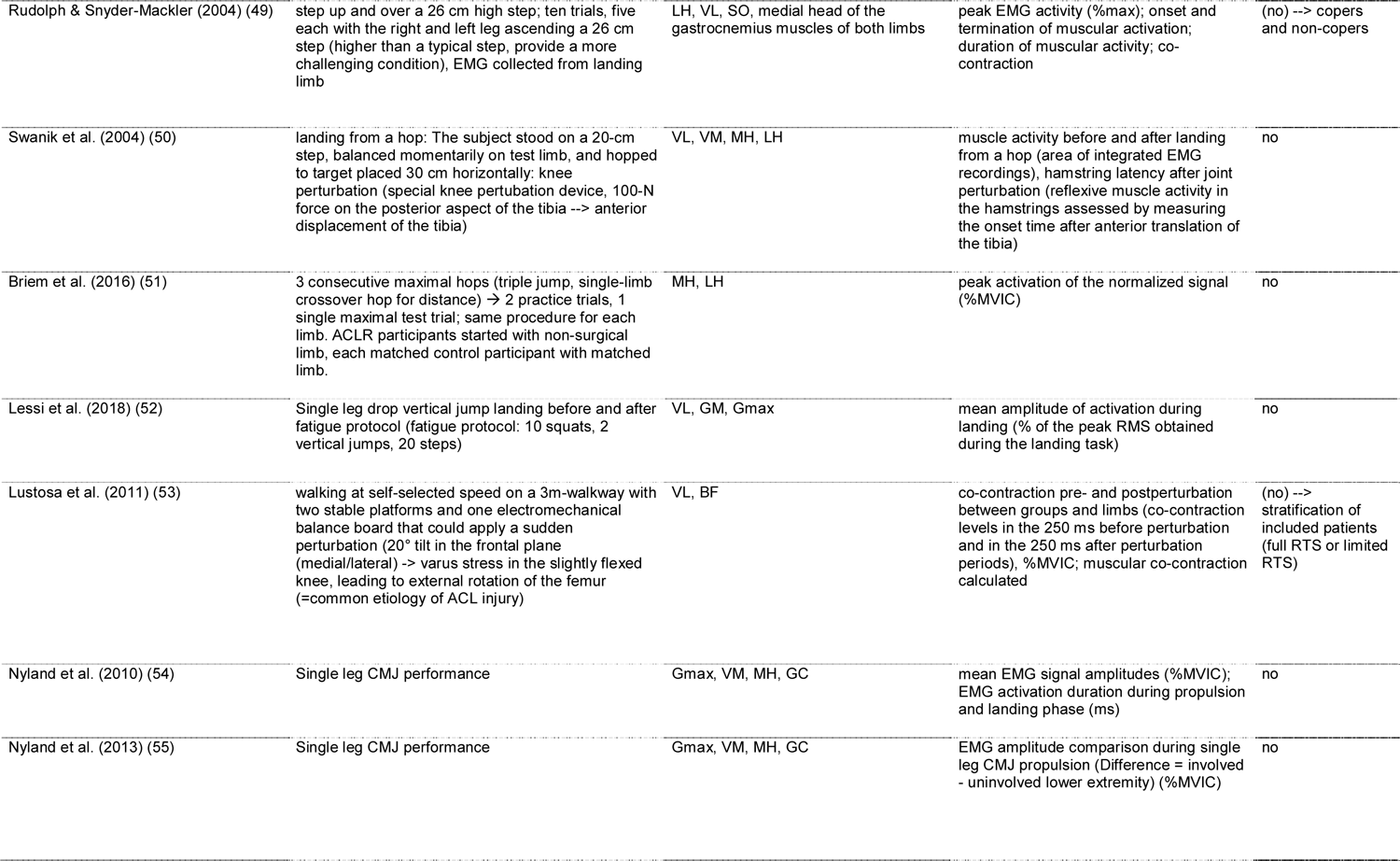
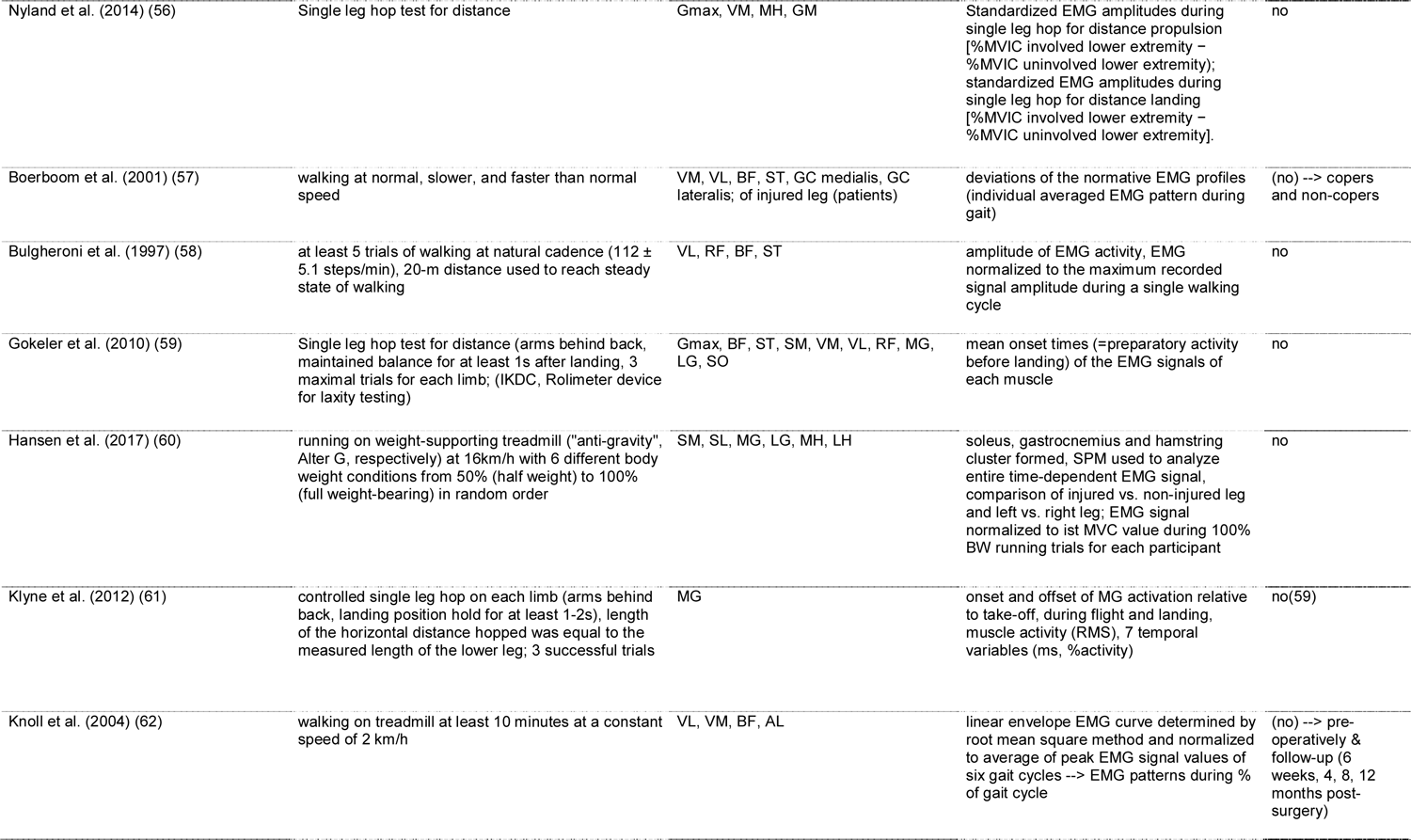
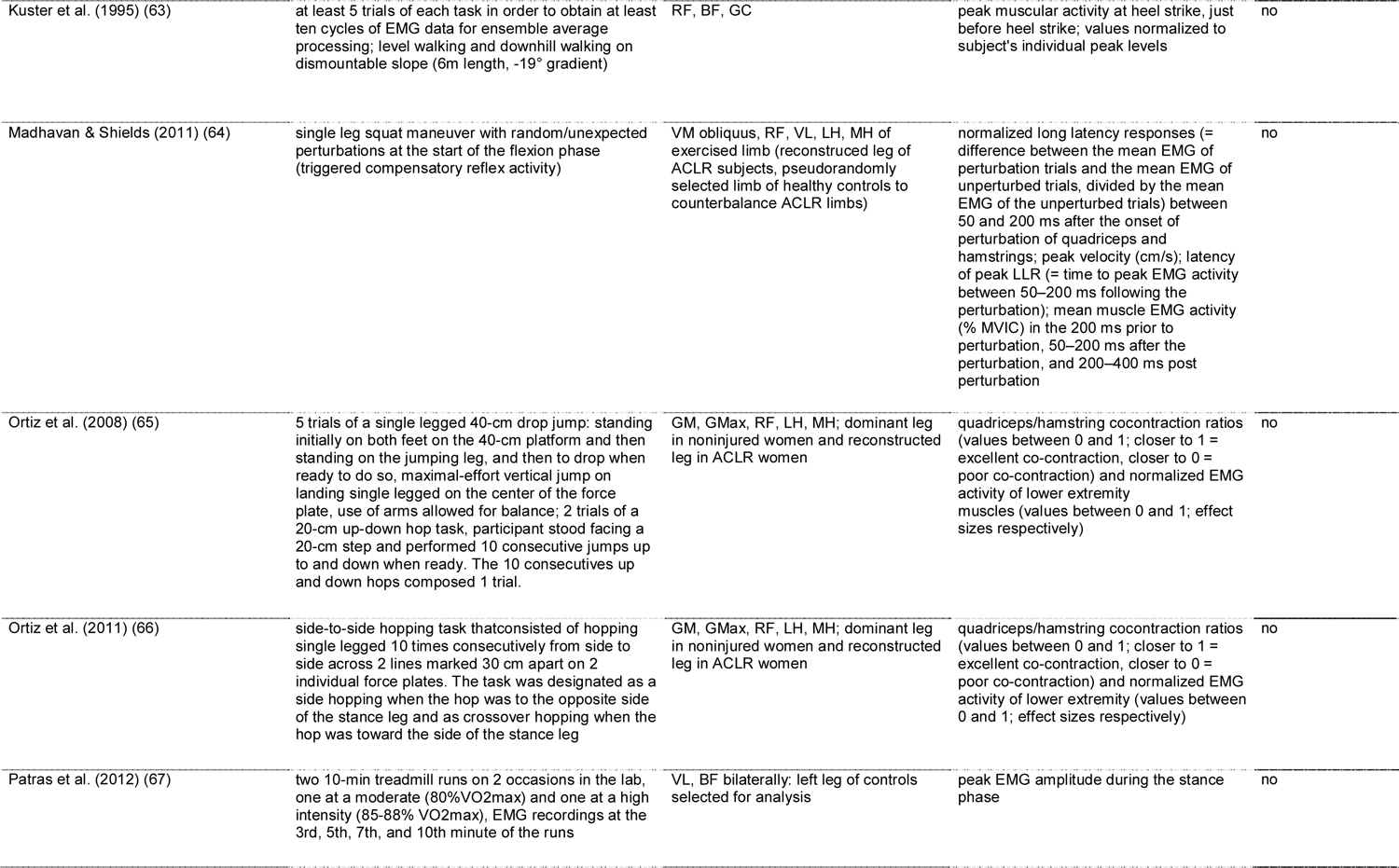
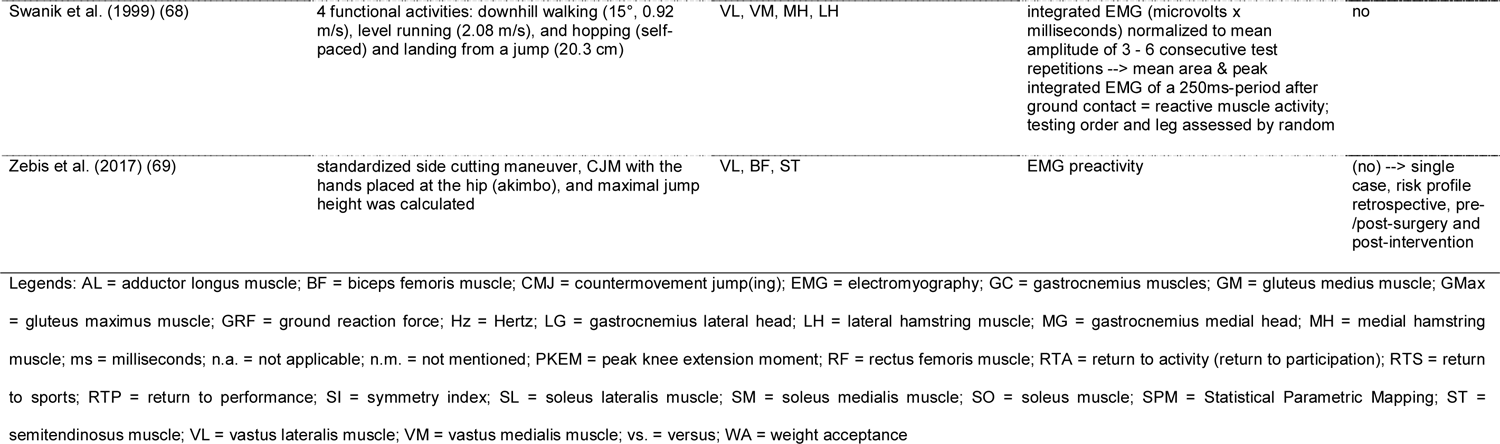
Characteristics of methods of included studies

#### Interventions

The number of muscles assessed ranged from one (37, 45, 61) to ten (59). Mainly muscle activity of four muscles of the thigh, vastus lateralis, vastus medialis, biceps femoris and semitendinosus, had been assessed. However, there were also studies measuring the adductor longus (39, 62), gluteus medius (39, 65, 66), gluteus maximus (41, 52, 54–56, 59, 65, 66), and calf muscles such as soleus, medial and lateral gastrocnemius (47-49, 54-56, 59, 60, 63).

The tasks used were very diverse: there were activities of daily life such as walking on even ground and downhill (33, 47, 53, 57, 58, 62, 63, 68), and stair climbing (13, 49). Other activities went more towards sports such as running (44, 45, 60, 67, 68) and jumping (36, 37, 39–42, 48, 50–52, 54–56, 59, 61, 65, 66, 68) where mainly the single-leg hop for distance, drop jumps and countermovement jumps were used. Some authors chose typical rehabilitation exercises such as forward lunges (34), Nordic hamstrings or hamstrings curls (35) and squats (64). At the other end of the scale, more complex, highly demanding, sport-specific tasks such as an instep soccer kick (38) or a sidecutting maneuver (69) were reported. Only few research groups used perturbation platforms to simulate injury mechanisms during walking (53) or squatting (46, 64), or applied devices to stress the ACL in the posterior-anterior direction (50). In addition, two studies even investigated the influence of fatigue on neuromuscular control (41, 52).

#### Outcomes

All included studies used surface EMG as method to assess neuromuscular control and provided EMG-related variables such as peak and mean amplitudes, timing and peak of muscle activity, preparatory and reactive muscle activity, on- and offset of muscular activation, co-activation/co-contraction ratios, or asymmetry index. The outcome variables were expressed as percentage of maximum voluntary (isometric) contraction (%MVIC or %MVC) or reported in microvolts or milliseconds according to the variable chosen in amplitude or time domain.

### Decision for Return to Sports RTS

None of the included studies used the surface EMG measurements to decide upon readiness for RTS (Table 4). However, the results from about a third of the studies (31.6%, 12 studies) could provide useful information by the choice of the assessed groups such as copers versus non-copers (33, 34, 47–49, 57), intervention and control group from the same team or level/league (35, 38, 40), data from pre-injury/pre-surgery including post-surgical follow up (62, 69) or participants with full RTS versus limited RTS (53). In addition, two studies even investigated the influence of fatigue on neuromuscular control (41, 52).

## Discussion

The aim of this systematic review was to summarize the scientific literature regarding EMG-related assessments for neuromuscular control in patients with an ACL injury (either treated surgically or conservatively). The second aim was to analyze whether these assessments for neuromuscular control were used to decide upon readiness for RTS in these patients.

There were many factors present which could have an influence on neuromuscular control:

### Influence by type of comparison (intra-versus inter-subject)

The use of the contralateral, non-injured leg in intra-subject comparison, without a “real” control group (42, 44) may lead to an overestimation of the physical performance in the ACL reconstructed or -injured leg. After ACLR, functional performance is often expressed with the LSI (70). As the non-affected limb may also have deteriorated, the LSI may overestimate the right time for a safe RTS, and therefore, the risk for secondary injury may be higher (23). In acutely injured ACL patients, intra-individual comparison showed bilateral consequences during stair ascent and indicates an alteration in the motor program (‘‘pre-programmed activity’’) (71). In addition, in case of a case-controlled study design, the subjects in the control group should be matched to the ACL participants regarding sex, age, body mass, height, activity level and leg dominance.

### Influence by level of activity and fatigue

Some of the included studies used very challenging, sports-specific task to assess neuromuscular control, some even assessed neuromuscular control after fatiguing tasks. It is known that most of ACL tears are non-contact injuries happening at the end of a training session or a play (72). Therefore, the closer the task to the sports and injury-risky situation, the safer the decision towards full RTS or even return to competition will be. However, assessing performance-based tests or movement quality may be more difficult to standardize, require more complex equipment and large amounts of space. But if only impairments will be tested, there will be a lack of information regarding an “athlete’s capacity to cope with the physical and mental demands of playing sport” (73). It is therefore recommended to search for a standardized assessment close to the injury mechanism.

### Influence by gender

Not all included studies reported findings of mixed groups separately by gender. Some did not even state whether study participants were male or female. This could partly be explained by the date of publication as gender difference in ACL patients has not been in the focus of former ACL research. It is known that female athletes are more likely to sustain an ACL injury than men (74, 75); the increased risk is probably multifactorial (76). Several studies indicate that hormonal factors play a role (3, 77) contributing to an increased laxity of ligaments in the first half of the menstrual cycle. However, biomechanical and neuromuscular aspects as indicators are discussed controversially in literature: Gender-specific neuromuscular adaptations and biomechanical landing techniques are considered being the most important ones to explain the increased risk of injury in women (78, 79). The higher risk for females to suffer from an ACL injury can be explained by motion and loading of the knee joint during performance (74). Female athletes typically perform movements in sports with a greater knee valgus angle than men. Therefore, the amount of stress on the ACL in these situations is higher caused by a high activation of the quadriceps despite limited knee and hip flexion, greater hip adduction and a large knee adduction moment (80, 81). The dominance of the quadriceps muscle in women could contribute to increased anterior tibial translation (82, 83) and was found in various activities such as jumps and cutting maneuvers (84–86). Moreover, females typically land with an internally or externally rotated tibia (87), leading to an increased knee valgus stress due to greater and more laterally orientated ground reaction forces (83). In contrast, other researchers did not find any gender-specific differences in the quadriceps-hamstrings ratio (88), not even in landing and cutting maneuvers (89). A systematic review summarized biomechanical gender differences and stated that these were based on questionable clinical relevance (89). In addition, strength-paired women and men showed no significant differences in neuromuscular activity (90).

### Influence by treatment

The included studies reported different treatment options (ACLR with different graft types, conservative treatment). Depending on the classification of the participants in copers and non-copers, the results in neuromuscular control may differ from a population of ACLR participants. Therefore, all researchers who worked with copers and non-copers made intra- and inter-group comparisons without an ACLR group. A Cochrane review revealed low evidence for no difference in young, active adults after two and five years after the injury, assessed with patient-reported outcomes. However, many participants described as “non-copers” with unstable knee with conservative treatments remain symptomatic, and therefore, later opt for ACL surgery (91). It has been described that persistent co-contraction and joint stiffening in these “non-copers” is likely to be due to an abnormal neuromuscular strategy failing to restore joint stability in these ACL deficient group (92). Furthermore, the choice of graft would influence the neuromuscular control of measured muscles due to the morbidity of the harvesting site of the graft (e.g. hamstrings).

### EMG variables

If the researchers mentioned the procedures for collecting EMG data, they referred to standardized applications and guidelines such as SENIAM (Surface ElectroMyoGraphy for the Non-Invasive Assessment of Muscles) (93). The provided EMG-related variables were in accordance to the ones mentioned in a systematic review searching for knee muscle activity in ACL deficient patients and healthy controls during gait (14). Current literature suggests greater co-contraction indices, increased joint stiffness and earlier muscle activation onset times as measures of neuromuscular function reflecting the incomplete restoration of normal joint stability (19, 92). Some of the included studies reported values of muscle onset activity in milliseconds and percentage of gait cycle as a systematic review did by summarizing and quantitatively analyzing muscle onset activity prior to landing in patients after ACL injury (20). However, no cut-off values out of EMG-related variables were provided to determine an adequate level of neuromuscular control. Moreover, some of the researchers only provided integrated EMG values which would make it difficult to be compared to other studies using the respective units (milliseconds, millivolts) or widely used percentage values (%MVIC, %MVC).

### Return to sports (RTS)

Regarding the determination of RTS after ACLR, there is some evidence for the use of functional performance tests, which had also been widely used in the included studies. Multiple functional performance measures – a battery including strength and hop tests, quality of movement and psychological tests (25) - might be more useful for the determination of RTS than a single performance measure. However, it is still unclear, which measures should be used to bring athletes safely back to RTS with a low risk of a second ACL injury (25). Currently used RTS criteria or assessments, such as time, strength tests, hop tests, patient-reports, clinical examination, thigh circumference, ligamentous stability, range of motion, effusion and performance-based criteria, may be suboptimal at reducing the risk of a second ACL injury (73, 94). Recovery of neuromuscular function was mentioned to be important because of the existing connection between the variables time since surgery and the risk for re-injury of the knee joint; but adequate assessment procedures to assess neuromuscular function are still a matter of debate (7). In contrast, authors of an included study stated that “studies like ours that focus on the objective measurement of the change of the muscle latency time over time may allow patients to return to full activity and to sports earlier than the standard time of 6–12 months” (42). However, this statement only based on one outcome measure and contrasts with current criterion- and time-based recommendations for RTS. Therefore, this recommendation seems to be rather dangerous.

### Limitations

The sample size of all the studies was quite low, however, providing reasonable sample size calculations and depending on the variable investigated, the results were acceptable. Furthermore, the more restrictive the inclusion criteria for the participants, the more homogeneous the intervention and the control groups were, but the more challenging the recruitment process was, leading to smaller groups to be investigated.

The used assessment for the risk of bias, the Downs and Black checklist (31) in a modified form (29, 32) is designed for randomized and non-randomized controlled studies, however, the latter score lower in some items, get lower total scores and therefore a worse overall rating of the methodological quality. Despite this disadvantage, we decided to use the modified checklist as we could assess all studies with different designs included in this systematic review. However, the use of total scores and choice of cut-off values for low, medium and high risk of bias, respectively, were arbitrary and not based on literature.

## Conclusions

### Implications for clinical practice

This systematic review summarized assessments using EMG variables for neuromuscular control of the knee in patients suffering from an ACL injury (either treated surgically or conservatively). Despite 38 articles providing a wide range of EMG-related assessments, none was used to decide upon readiness towards a safe and successful RTS in patients after an ACL injury. So far, there is no diagnostic measure to assess neuromuscular control and therefore, clinicians should use a multimodal approach including assessments for active and passive knee stability under different sports-related conditions but be aware of not being able to evaluate neuromuscular control in depth without EMG-related assessments. Moreover, the widely used LSI may overestimate the physical performance of an ACL patient as the non-affected limb is likely to have deteriorated, too.

### Implications for further research

Additional studies are needed to define readiness towards RTS by assessing neuromuscular control in adult ACL patients with EMG. Further research should aim at finding reliable and valid, EMG-related variables to be used as diagnostic tool for neuromuscular control. Due to the heterogeneity in participants, interventions and outcomes used, future studies should aim at more homogenous patient groups, evaluate females and males separately, provide adequately matched healthy subjects (gender, height, weight, activity level etc.), control for confounding factors such as type of treatment, and use tasks close to the injury mechanism, as sport specific as possible, respectively. Moreover, it would be interesting to assess not only lower leg but pelvic and core muscles in addition. This would help to give insight in the complex field of ACL injuries and subsequent rehabilitation strategies, and therefore improve knowledge towards a safe RTS in these patients.

Appendix A_search string

Appendix B_Risk of bias assessment

## Declarations

### Ethics approval and consent to participate

Not applicable.

### Consent for publication

Not applicable.

### Availability of data and material

The datasets used and analyzed in the current study are available from the corresponding author on reasonable request.

## Authors’ contributions

AB participated in the design of the study, contributed to data collection/reduction/analysis and interpretation of results and was the main contributor in writing the manuscript; IK contributed to data collection, reduction and analysis; IB and DV participated in the design of the study; HB participated in the design of the study and was an important contributor in writing the manuscript, contributed to data analysis and interpretation of results. All authors contributed to the manuscript writing, read, and approved the final version of the manuscript and agreed with the order of authors as listed.

## Data Availability

All data referred to in the manuscript are available from the first author upon request.

## Acknowledgments

We would like to thank to all the following people for their assistance and contribution regarding this work: the librarians of the Bern University (Department of Health Professions) and the University of Bern (Institute for Sports Science) for their support before and during the literature search, M. Akter for her help in extracting relevant key words and A. Busch for her critical review of the proposal and support to refine the search strategy.

## List of abbreviations

ACL: anterior cruciate ligament

ACLR: anterior cruciate ligament reconstruction

EMG: electromyography

LSI: Limb Symmetry Index

PEDro: Physiotherapy Evidence Database

PICOS: Participants-Intervention-Control-Outcome-Study design

PRISMA: Preferred Reporting of Items for Systematic Reviews and Meta-Analyses

PROSPERO: International Prospective Register of Systematic Reviews

RTS: return to sports

SENIAM: Surface ElectroMyoGraphy for the Non-Invasive Assessment of Muscles

TAS: Tegner Activity Score

## Authors’ information

AB, MPTSc, MSc in Physiotherapy, PT, is Deputy Head of the research group “Neuromuscular control” and Co-Leader of the Bachelor of Science in Physiotherapy at the Bern University of Applied Sciences, Department of Health Professions, Division of Physiotherapy in Bern (Switzerland). AB is well-trained in orthopedics, sports physiotherapy and active rehabilitation in different settings. She works as lecturer and researcher and does her doctoral studies at the University of Antwerp (Belgium) in the field of neuromuscular control and ACL injuries.

IK, PhD, MSc in Physiotherapy, M.A. in Adult Education, PT, is Head of the research group “Pelvic Floor and Continence” and Co-Leader of the Bachelor of Science in Physiotherapy at the Bern University of Applied Sciences, Department of Health Professions, Division of Physiotherapy in Bern (Switzerland). She is well-trained in pelvic floor rehabilitation, EMG assessments and analyses with different methods.

IB, PhD, PT, works as tenure track lecturer at the University of Antwerp, Faculty of Medicine and Health Sciences, Department of Rehabilitation Sciences and Physiotherapy in Wilrijk (Belgium) in the Bachelor and Master study programs. Her main interests regarding research are in the field of musculoskeletal diseases (lower extremity and lumbopelvic region), assessments in physiotherapy and pain management.

HB, Professor, PhD in Sport and Movement Science, is Head of Applied Research and Development and Head of the research group “Neuromuscular control” at the Bern University of Applied Sciences, Department of Health Professions, Division of Physiotherapy in Bern (Switzerland). His main interests in research are development of diagnostic and interventional assessments for musculoskeletal problems, influence of neuromuscular control by external (training/exercise, therapy, aids) and internal factors (gender, age, etc.) and development of quality control measures in physiotherapy.

DV, Professor, PhD, PT, is senior lecturer at the at the University of Antwerp, Faculty of Medicine and Health Sciences, Department of Rehabilitation Sciences and Physiotherapy in Wilrijk (Belgium). He has expertise in sports-related injuries, physical conditioning (muscle strength, motor control exercises), technology use in the field of physiotherapy, exercise therapy and rehabilitation in different pathologies. DV is first or co-author of several systematic reviews and meta-analysis.

