## Appendix A_search string for "Which assessments are used to analyze neuromuscular control by electromyography after an anterior cruciate ligament injury to determine readiness to return to sports? A systematic review"

**Belonging to the manuscript entitled**

**Appendix A: Search string for MEDLINE/PubMed**

((((((((anterior cruciate ligament[MeSH Terms]) OR anterior cruciate ligament[Title/Abstract]) OR anterior cruciate ligaments[Title/Abstract]) OR anterior cranial cruciate ligament[Title/Abstract]) OR anterior cranial cruciate ligaments[Title/Abstract]) OR ACL[Title/Abstract])) AND (((((((((((((((((((anterior cruciate ligament injuries[MeSH Terms]) OR anterior cruciate ligament injuries[Title/Abstract]) OR anterior cruciate ligament injury[Title/Abstract]) OR rupture[Title/Abstract]) OR ruptures[Title/Abstract]) OR strain[Title/Abstract]) OR strains[Title/Abstract]) OR sprain[Title/Abstract]) OR sprains[Title/Abstract]) OR tear[Title/Abstract]) OR tears[Title/Abstract]) OR ((strains and sprains[MeSH Terms]))) OR ((strains[Title/Abstract] AND sprains[Title/Abstract]))) OR injury[Title/Abstract]) OR injuries[Title/Abstract]) OR partial tear[Title/Abstract]) OR partial tears[Title/Abstract]) OR deficiency[Title/Abstract]))) AND (((((((((((((((((anterior ligament reconstruction[MeSH Terms]) OR anterior ligament reconstruction[Title/Abstract]) OR anterior ligament reconstructions[Title/Abstract]) OR anterior cruciate ligament/surgery[Title/Abstract]) OR anterior cruciate ligament/surgery[MeSH Terms]) OR anterior cruciate ligament/surgery[Title/Abstract]) OR reconstructive surgical procedures[MeSH Terms]) OR reconstructive surgical procedures[Title/Abstract]) OR reconstructive surgical procedure[Title/Abstract]) OR reconstruction[Title/Abstract]) OR reconstructions[Title/Abstract]) OR reconstructive[Title/Abstract])))) AND (((((((((neuromuscular control[Title/Abstract]) OR neuromuscular activity[Title/Abstract]) OR sensorimotor control[Title/Abstract]) OR muscle activity[Title/Abstract]) OR muscular activity[Title/Abstract]) OR active stability[Title/Abstract]) OR active joint stability[Title/Abstract]) OR active knee stability[Title/Abstract]) OR active knee joint stability[Title/Abstract]))) AND ((((((((((((((((electromyography[MeSH Terms]) OR electromyography[Title/Abstract]) OR surface electromyography[Title/Abstract]) OR electromyogram[Title/Abstract]) OR EMG[Title/Abstract]) OR amplitude[Title/Abstract]) OR timing[Title/Abstract]) OR mean activity[Title/Abstract]) OR peak activity[Title/Abstract]) OR duration of activity[Title/Abstract]) OR onset of activity[Title/Abstract]) OR offset of activity[Title/Abstract]) OR on-off-pattern[Title/Abstract]) OR pre-activity[Title/Abstract]) OR latency[Title/Abstract]) OR reflex response[Title/Abstract])
