## Appendix B_Risk of bias assessment for "Which assessments are used to analyze neuromuscular control by electromyography after an anterior cruciate ligament injury to determine readiness to return to sports? A systematic review"

**Belonging to the manuscript entitled**

Methodological quality assessment from Downs & Black (1998) (31), modified by Ramsey et al. (2019) (29) and Pairot-de-Fontenay et al. (2019) (32) with adaptations

| **Category** | **Question number in Downs and Black** | **Question** | **Application to this review** |
| --- | --- | --- | --- |
| Reporting | 1. | Is the hypothesis/aim/objective of the study clearly described? | Score of  1 = if hypothesis/aim/objective described.  0 = for NO description. |
| Reporting | 2. | Are the main outcomes to be measured clearly described in the introduction or methods section? | Score of  1 = if main outcome measure(s) described in introduction or methods.  0 = for NO description. |
| Reporting | 3. | Are the characteristics of the patients included in the study clearly described? | Score of “1” for YES  “0” for NO defined criteria.  ACLR group: Clear inclusion and exclusion criteria; Injured leg: primary  ACLR, non-injured leg: history of surgery, knee injury or pathology; Control group: history of surgery, knee injury or pathology. |
| Reporting | 4. | Are the interventions of interest clearly described? | assessment / measurement of lower extremity of ACL injured people or their controls to assess neuromuscular control of the knee/active knee stability/knee joint stability under dynamic conditions  Score of “1” if intervention clearly described. “0” for no or insufficient/unclear description.  “X” for unable to determine |
| Reporting | 5. | Are the distributions of principal confounders for each group to be compared clearly described? | Age, gender, BMI or body mass & height respectively, activity level/sports activity (Tegner score desirable, but not mandatory) were considered the main confounders. If >1 group of patients: time since injury as additional confounder  If 3 or 4 of these were specified: score of “2” If 1 or 2 of these were specified: score “1”  If none of these were specified: score “0” |
| Reporting | 6. | Are the main findings of the study clearly described? | Score of “1” if YES, main findings clearly described respectively. Score  “0” for NO description. |
| Reporting | 7. | Does the study provide estimates of the random variability in the data for | SD, SE, CI were considered for measures of variability. Score of “1” if any of these measures of variability given. Score “0” for no description |

|  |  | the main outcomes? | of measures of variability.  In non-normally distributed data: inter-quartile range of results should be reported; in normally distributed data: SE, SD or CI should be  reported |
| --- | --- | --- | --- |
| Reporting | 10. | Have actual probability values been reported (e.g. 0.035 rather than <0.05) for the main outcomes except where probability value is less than 0.001? | Score of “1” if actual probability values described. Score “0” for no description. |
| External validity bias | 11. | Were the subjects asked to participate in the study representative of the entire population from which they were recruited? | Score of “1” if participants recruited were from the community. Score “X” for unable to determine and “0” if the participants were from a single hospital. |
| External validity bias | 12. | Were those subjects who were prepared to participate representative of the entire population from which they were recruited? | Score of “1” if participants who contacted were from the community and the cohort was finalised from this community-based population based on the inclusion criteria. Score “0” for no description.  “X” for unable to determine |
| External validity bias | 13. | Were the staff, places, and facilities where the patients/participants were treated, representative of the treatment/intervention the majority of patients received? | Score of “1” if intervention/task was representative of that in use in the source population 🡪 stairs, gait, walking, running, jumping, squats, stop & go manoeuvres etc. Score “0” if e.g. intervention was undertaken in a specialist centre unrepresentative of the locations/hospitals/private practices most of the source population would attend 🡪 activities in a movement lab or similar tasks such as treadmill running or walking, legpress, dynamometry etc.  “X” for unable to determine |
| Internal validity bias | 14. | Was an attempt made to blind study subjects to the intervention they have received? | Score of “1” if an attempt was made or the study participant was blinded to the intervention. Score “0” if no attempt was made despite the possibility to blind participants. Score “X” if unable to determine, study design was not a RCT respectively. |
| Internal validity bias | 15. | Was an attempt made to blind those measuring main outcomes of the intervention? | Score of “1” if YES, given if blinding done during data processing. Score “0” for NO description. Score “X” if unable to determine, study design was not a RCT respectively. |
| Internal validity bias | 16. | If any of the results of the study were based on “data dredging”, was this made clear? | Score of “1” for clearly mentioning the outcome measures planned. Score “0” if data dredging was there.  “X” for unable to determine |

| Internal validity bias | 17. | In trials and cohort studies, do the analyses adjust for different lengths of follow-up of patients, or in case-control studies, is the time period between the intervention and outcome the same for cases and controls? | Score of “1” if YES. Score “0” if NO. Score “X” if unable to determine, in case of a healthy control group respectively. |
| --- | --- | --- | --- |
| Internal validity bias | 18. | Were the statistical tests used to assess the main outcomes appropriate? | Score of “1” if YES, appropriate statistical tests used. Score “0” for non- appropriate statistical tests and “0” for no description.  “X” for unable to determine |
| Internal validity bias | 20. | Were the main outcome measures used accurate (valid and reliable)? | Score of “1” if reference given for reliability or validity of the outcome measures used or use of an established measure/assessment such as EMG. Score “0” for no description, use of a new, not sufficiently tested measure.  “X” for unable to determine |
| Internal validity: selection bias | 21. | Were the patients in different intervention groups (trials and cohort studies) or were the cases and controls (case-control studies) recruited from the same population? | It was considered important that groups were matched for age (within a mean of 5 years), BMI or body mass & height respectively, sport level, gender. Score of “1” if age, BMI or body mass & height respectively, sport level and gender matched in both groups. In other words: no statistical group differences. Score “0” if groups were not matched and “0” for no description. Score “X” if unable to determine (cohort studies, case-control studies). Apply “IC” when comparison with the  contralateral limb was done. |
| Internal validity: selection bias | 22. | Were study subjects in different intervention groups (trials and cohort studies) or were the cases and controls (case-control studies) recruited over the same period of time? | Score “1” if YES. Score “0” if NO. Score “X” if unable to determine, in case of cross-sectional study, time not specified. |
| Internal validity: selection bias | 23. | Were study subjects randomised to intervention groups? | Score “1” if YES. Score “0” if NO. Score “X” if it is another study design than a RCT, e.g. a case-control study or if unable to determine. |
| Internal validity:  selection | 24. | Was the randomised intervention assignment concealed from both patients and health care staff until recruitment was complete and irrevocable? | Score “1” if YES. Score “0” if NO. Score “X” if it is another study design than a RCT, e.g. a case-control study or if unable to determine. |

| bias |  |  |  |
| --- | --- | --- | --- |
| Internal validity: selection bias | 25. | Was there adequate adjustment for the confounding in the analysis from which the findings were drawn? | A score of “1” was applied If neuromuscular control/active knee stability was not significantly different between groups, alternatively if this was considered a confounding factor in the statistical analysis.  Score of “0” if neuromuscular control/active knee stability was significantly different between groups and not considered as a confounding factor in the statistical analysis. Score of “X” for unable to determine. Apply “IC” when comparison with the contralateral limb  was done. |
| Internal validity: selection bias | 26. | Were losses of patients to follow-up taken into account? | Score “1” if YES. Score “0” if NO. Score “X” if unable to determine. Score “X” if e.g. study design without follow-up such as cross-sectional or cohort study |
| Power | 27. | Were appropriate power calculations reported? | A score of “1” was applied when a power or a sample size calculation was provided. If these were not given or there was no explanation whether the number of participants was appropriate a “0” was applied. |
| Explanations/Abbreviations: ACLR: Anterior cruciate ligament reconstruction; BMI: Body Mass Index; CI: Confidence interval; SD: Standard deviation; SE: Standard error; IC: intrasubject comparison (comparison with contralateral limb) | | | |
